# A scoping literature review of global dengue age-stratified seroprevalence data: estimating dengue force of infection in endemic countries

**DOI:** 10.1101/2023.04.07.23288290

**Authors:** Anna Vicco, Clare McCormack, Belen Pedrique, Isabela Ribeiro, Gathsaurie Neelika Malavige, Ilaria Dorigatti

## Abstract

Dengue poses a significant burden worldwide, and a more comprehensive understanding of the heterogeneity in the intensity of dengue transmission within endemic countries is necessary to evaluate the potential impact of public health interventions.

This scoping literature review aimed to update a previous study of dengue transmission intensity by collating global age-stratified dengue seroprevalence data published in the Medline and Embase databases from 2014 to 2022. These data were then utilized to calibrate catalytic models and estimate the force of infection (FOI), which is the yearly per-capita risk of infection for a typical susceptible individual.

We found a total of 44 new publications containing 47 relevant datasets across 20 endemic countries. Together with the previously available average FOI estimates, there are now 280 dengue average FOI estimates obtained from seroprevalence data and 149 estimates obtained from case-notification data available across the world.

The results showed large heterogeneities in average dengue FOI both across and within countries. These new estimates can be used to inform ongoing modelling efforts to improve our understanding of the drivers of heterogeneity in dengue transmission globally, which in turn can help inform the optimal implementation of public health interventions.

**Author summary:** In this work, we conducted a scoping literature review to collate global dengue age-specific seroprevalence data from dengue endemic areas published between 2014 and 2022. These data were used to calibrate mathematical models and estimate the average yearly force of infection (FOI), which is a fundamental measure of transmission intensity. FOI estimates can be used to quantify the risk of infection, disease burden and the potential impact of new interventions, such as vaccination.

In addition, the FOI estimates generated in this study contribute to ongoing efforts to better characterise and map dengue transmission intensity worldwide.

## Introduction

Dengue is a rapidly spreading mosquito-borne viral infection transmitted to human by *Aedes* mosquitoes, primarily affecting tropical and sub-tropical regions [1]. With half of the world’s population at risk of contracting the disease, dengue is the leading arboviral disease among humans. However, its transmission dynamics in several countries remain poorly understood [2].

Notifications of suspected or virologically confirmed dengue cases, together with information on the age and location of the reported cases, provide the necessary information to reconstruct the immunity profile (i.e., the age-dependent susceptibility to infection) of a local population. This, in turn, can be used to estimate the transmission intensity of dengue, as defined by the force of infection (FOI), which is the per-capita rate of infection for a susceptible individual. However, while routinely collected case-notification data exists for several locations in South East Asia and Latin America, case reporting in Africa is more variable and not mandatory due to the limited availability of laboratory diagnostic tools, and the lack of point-of-care diagnostic – with cases mainly reported in sporadic outbreaks or in individual case reports - which has hindered arbovirus surveillance across the continent [3].

Serological surveillance involves the collection and testing of blood samples for the presence of IgG antibodies against dengue virus using qualitative or quantitative serological assays. This approach allows for the detection of previous infections, regardless of the level of symptoms experienced upon infection [4]. This is particularly important for diseases such as dengue, where asymptomatic infections are very frequent [5] and antibody levels against the infecting serotype are long-lived. By providing information on past exposure, serological surveillance is particularly useful for gaining insight into the historical circulation of dengue and for reconstructing the immunity profile of a population. This, in turn, can be used to inform surveillance, preparedness planning, the potential implementation of existing and new vector control strategies, such as the release of Wolbachia-carrying mosquitoes [6], and the optimal implementation of vaccination programs [7,8]. Specifically, age-stratified serological surveys provide key data on the immunity profile of the population, which allows us to estimate the FOI.

Several different approaches have been taken to map the risk of dengue infection in the current and changing climate [9], using multiple risk metrics including environmental, climate and habitat suitability [2,10,11], R0 estimates [12] and occurrence data [13]. While more limited than occurrence data, FOI estimates have the advantage of allowing us to estimate changes in the burden of infection and disease in the presence of large-scale interventions, such as age-targeted vaccination campaigns in endemic settings, where accounting for pre-existing immunity is important [14]. The first global map of dengue FOI was developed by Cattarino et al. [15], where machine learning was used to link 382 geolocated dengue force of infection (FOI) estimates with a set of ecological and demographic variables. Dengue FOI predictions were then imputed globally, according to underlying climatic, environmental and demographic conditions, including in places with no (serological nor case-based) dengue surveillance. Ongoing efforts aiming to validate and refine the FOI projections and burden estimates obtained by Cattarino et al. require an update of an earlier literature review, conducted by Imai et al. in 2014 [4], which provided the serologically-derived FOI estimates for the model developed by Cattarino et al. [15].

In this work, a literature search was conducted to collect global age-stratified seroprevalence data on dengue published since 2014. The objective of this literature search was to update current knowledge of the transmission intensity of dengue in endemic countries. As in Imai et al. [4], these data were then used to calibrate catalytic models to estimate the setting-specific dengue force of infection (FOI). These new estimates are crucial for updating live datasets and for quantifying the extent of dengue transmission in the last decade. Furthermore, they play an important role in informing public health efforts to better characterise the epidemiology of dengue and to control the spread of this disease.

## Materials and methods

### Literature search

We searched Medline and Embase for publications reporting age-stratified dengue seroprevalence datasets published between January 2014 and September 2022 to complement the literature search previously conducted by Imai et al [4]. We used a Boolean search query with search terms:

**dengue AND sero* AND (prevalence OR seroprevalence OR positiv* OR seropositiv*)**

The specific search terms for each database are reported in the Supplementary material (Table S1 and S2). After removing duplicate articles, articles in languages other than English, and non-article publications (e.g., conference posters and book chapters), we retained all remaining papers for evaluation of their potential relevance according to the titles and abstracts. The selected papers were evaluated for full-text eligibility, with the aim of collating data on age-specific dengue seroprevalence studies conducted in the general population in specific, geolocated endemic settings. Following Brady et al [16], we defined endemic countries those with “good” or “complete presence” of dengue.

Given our focus on estimating the susceptibility profile of the population and the FOI, we excluded papers where only the overall seroprevalence (i.e. not stratified by age) was reported, or where the study focused on hospital cases, clinical trials of antivirals, vaccine studies, or reported a secondary analysis of data published elsewhere (in which case, we included the original article presenting the primary data when the publication date fell within the search criteria). From the selected papers, we recorded the country and specific location of the survey (when available), the age range of the subjects, the number of age groups tested and the population sizes, as well as the date of the survey and the type of test used.

### Estimating the force of infection

We used catalytic models and a Bayesian inference approach to estimate dengue force of infection *λ* from serological data, as done by Imai et al. [4]. We used a simple catalytic model (model A), which assumes a constant infection hazard *λ* and no waning of immunity, to fit most cross-sectional serological (IgG) datasets, with the proportion seropositive in age group *a* (z(a)) given by Eq (1).

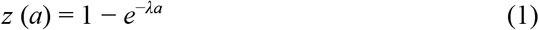

Whenever the data showed evidence of declining seroprevalence with age, we used model B, which is a modified version of model A with an extra parameter *α*, representing the antibody decay rate to account for antibody waning. Assuming a constant force of infection *λ* and decay rate *α*, the seroprevalence for model B is given by Eq (2).

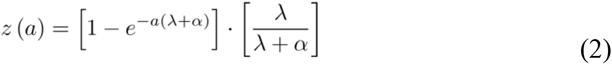

We fitted two versions of models A and B to the age-stratified serological data, assuming a binomial (models A1 and B1) and beta-binomial (models A2 and B2) distribution of the serological data, using the Hamiltonian Monte-Carlo algorithm implemented in the *CmdStanR* package [17,18]. We used Rhatt, n_eff_ and visual inspection of traceplots to assess convergence [17].

In models A2 and B2, we accounted for overdispersion of the data through an over-dispersion parameter *γ*, as described by Imai et al. [4]. The deviance information criterion (DIC) was used for model selection. For the datasets where we only had information on the percentage of people testing positive, and not on the total number of tested and testing people, we fitted the data using model C, where we assumed that the age-stratified seroprevalence (*X*_*a*_) was normally distributed with a mean (*µ*_a_ = *z*(*a*)) and a variance (*σ*) to be estimated, as described by Eq (3).

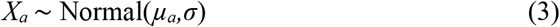

For each dataset we also report the average age of first infection, calculated as 1 / λ, and the age at which we expect x% seroprevalence (with x = 50% and 70%), calculated as − 1n(1− *x*)/*λ* for model A and as 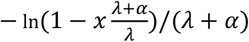 for model B. We calculated these statistics by extracting 100 random values from the posterior distribution of the parameters and reported its median and 95% CrI.

## Results

### Article selection and dataset characteristics

We found a total of 3,978 potentially relevant articles. Once we removed duplicates, 3,015 papers were retained for evaluation of titles and abstracts. Of these, 2,895 articles were found to be irrelevant for the purpose of the study and were therefore excluded. The remaining 120 papers were evaluated for full-text eligibility, from which we identified 44 studies reporting age-specific seroprevalence datasets (Fig 1) from surveys conducted between 2007 and 2022 in various dengue endemic countries (Fig 1).

**Fig 1.**
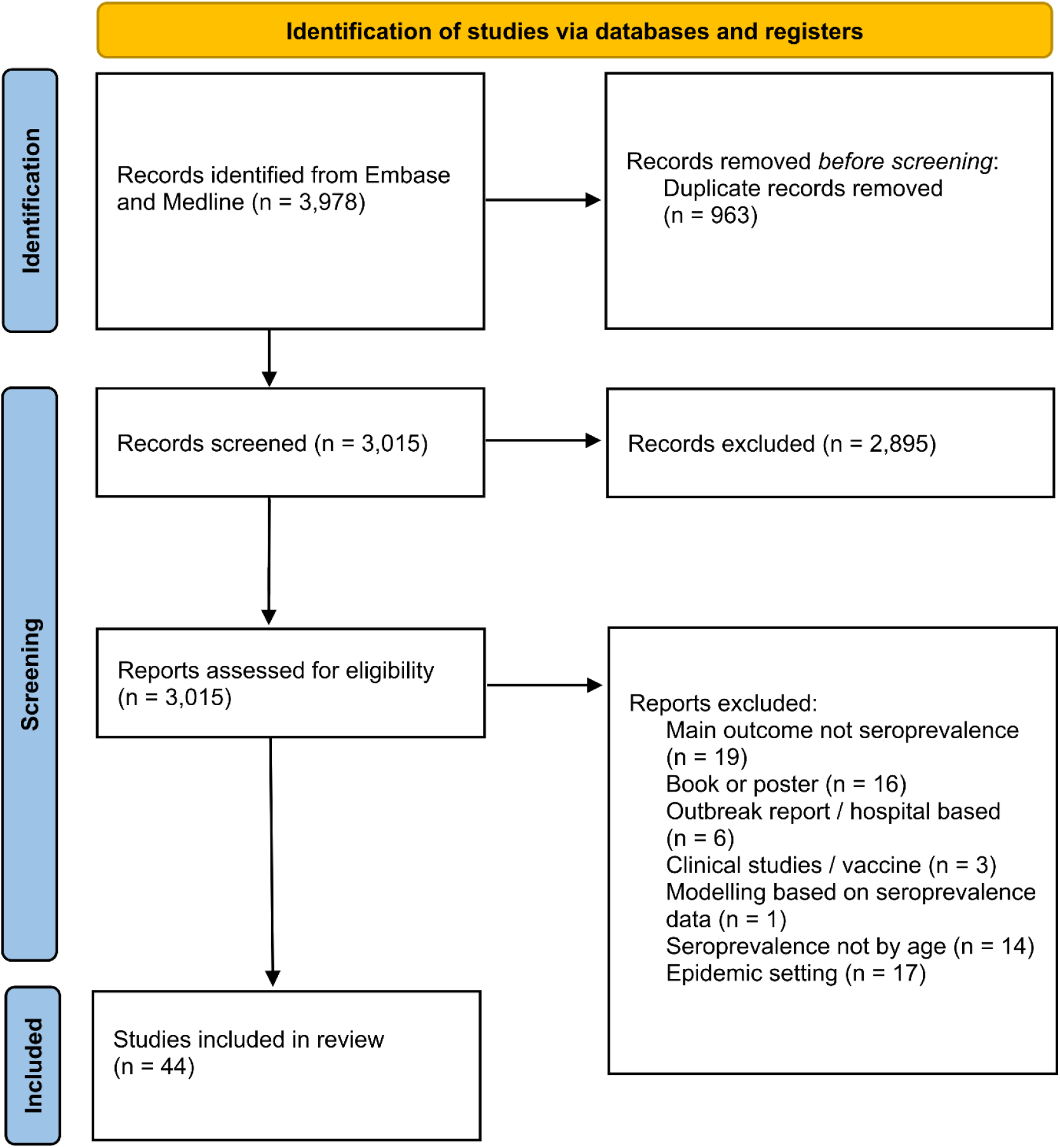
Workflow of the literature search. Literature review on dengue age-stratified seroprevalence from 2014 to the end of September 2022 in Embase and Medline using the word search “dengue AND sero* AND (prevalence OR seroprevalence OR positiv* OR seropositiv*)”.

Table 1 reports the main characteristics of each serosurvey, including the country, location, age range, number of age groups tested and size of the survey, type of test used and the date when the survey was performed. IgG ELISA was the assay most frequently used. Out of 44 studies, 11 focused on children and young people (up to 22 years old), mainly tested in schools and universities.

**Table 1.** Summary of age-specific seroprevalence datasets identified.

| Country | Location | Age range | Age groups | Total tested | Type of test | Ref. | Study date | Model | FOI (95% CrI) |
| --- | --- | --- | --- | --- | --- | --- | --- | --- | --- |
| Bangladesh | blood service | 18-67 | 5 | 507 | IgG ELISA | [19] | 2022 | B | 0.133 ( 0.038 , 0.288 ) |
| Bangladesh | Dhaka city | 0-67+ | 7 | 1,126 | IgG ELISA | [20] | 2012 | A | 0.045 (0.026, 0.071) |
| Brazil | Sau Paulo | 9-91 | 4 | 1,322 | IgG ELISA | [21] | 2015-2016 | A | 0.030 (0.013, 0.055) |
| Brazil | Fortaleza | 5-13 | 9 | 2,117 | indirect IgG ELISA | [22] | 2011-2015 | C | 0.096 (0.080, 0.115) |
| Burkina Faso | Ougadougou | 1-55 | 8 | 2,897 | indirect IgG ELISA | [23] | 2015-2017 | A | 0.063 ( 0.059 , 0.066 ) |
| Colombia | Anapoima, Apulo, Buenaventura, Quibdo, Tumaco, Tierralta | 4-95 | 4 | 1,318 | indirect IgG ELISA | [24] | 2013-2015 | A | 0.047 (0.020, 0.088) |
| Colombia | Quobdo | 0-45+ | 4 | 58 | indirect IgG ELISA | [25] | 2015* | A | 0.041 ( 0.028 , 0.060 ) |
| Ecuador | Town of Quininde | 0-60 | 11 | 115 | IgG ELISA | [26] | 2005-2009 | A | 0.094 ( 0.074 , 0.118 ) |
| FranceCaribbean | Guadeloupe and Martinique | 18-70 | 7 | 783 | IgG ELISA | [27] | 2011 | A | 0.078 ( 0.070 , 0.087 ) |
| Haiti | Gressier, Jacmel, Chabin | 0-50+ | 8 | 673 | indirect IgG ELISA | [28] | 2013 | B | 0.209 ( 0.100 , 0.352 ) |
| India | New Delhi, Hyderabad, Kalyani, Wardha, Mumbai, Bangalore | 05-9 | 6 | 2,556 | IgG ELISA | [29] | 2012 | A | 0.127 ( 0.121 , 0.134 ) |
| **India** | Pradesh, Tripura, Meghalaya, Assam, Bihar, West Bengal, Odisha, Rajasthan, Madhya Pradesh, Maharashtra, Andhra Pradesh, Karnataka, Tamil Nadu | 5-45 | 3 | 12,3 | indirect IgG ELISA | [30] | 2017 | A | 0.046 (0.017, 0.105) |
| **India** | Chennai | 5-40 | 5 | 1,01 | indirect IgG ELISA | [31] | 2011 | B | 0.072 ( 0.024 , 0.172 ) |
| **India** | Delhi | 0-50+ | 4 | 200 | capture IgG ELISA | [32] | 2012 | A | 0.043 ( 0.036 , 0.052 ) |
| **India** | Denpasar Bali | 0-40+ | 2 | 539 | IgG ELISA | [33] | 2020-2021 | A | 0.036 (0.011, 0.081) |
| **India** | Pune | 0-70+ | 13 | 1,434 | indirect IgG ELISA | [34] | 2017 | A | 0.078 ( 0.072 , 0.084 ) |
| **India** | Vadu area | 0-40+ | 5 | 702 | indirect IgG ELISA | [35] | 2011 | A | 0.021 ( 0.018 , 0.023 ) |
| **Indonesia** | national | 0-18 | 18 | 3,194 | IgG ELISA | [36] | 2014 | A | 0.144 ( 0.138 , 0.151 ) |
| **Indonesia** | national | 1-18 | 5 | 3,194 | IgG ELISA | [37] | 2014 | A | 0.149 ( 0.142 , 0.156 ) |
| **Kenya** | national | 15-64 | 3 | 1,091 | IgG ELISA | [38] | 2007 | A | 0.004 (0.004, 0.005) |
| **Malaysia** | Federal territory of Kuala Lumpur, Perak, Kedah, Penang, Johor, Pahang, Kelantan and Sabah | 7-18 | 4 | 1,417 | capture IgG ELISA | [39] | 2008-2009 | A | 0.010 (0.008, 0.012) |
| **Malaysia** | Damansara Damai | 0-25+ | 2 | 82 | IgG ELISA | [40] | 2018-2019 | A | 0.033 (0.010, 0.075) |
| **Malaysia** | Peninsular Malaysia | 35-74 | 4 | 1,417 | indirect IgG ELISA | [41] | 2006-2012 | A | 0.038 ( 0.036 , 0.041 ) |
| **Malaysia** | Petaling district | 0-60+ | 8 | 500 | IgG ELISA | [42] | 2018 | A | 0.059 ( 0.052 , 0.066) |
| **Malaysia** | Sungai Segamat | 18-55+ | 5 | 277 | capture IgG ELISA | [43] | 2015 | A | 0.029 ( 0.025 , 0.033 ) |
| **Mexico** | Pacific localities (Baja California, Baja California Sur, Nayarit, Sinloa, Sonora) | 6-17 | 3 | 271 | IgG ELISA | [44] | 2016 | C | 0.053 (0.013, 0.213) |
| **Mexico** | South-Central localities (Guerrero, Morelos, Oaxaca, Puebla, Veracruz) | 6-17 | 3 | 505 | IgG ELISA | [44] | 2016 | C | 0.061 (0.026, 0.190) |
| **Mexico** | South-East localities (Campeche, Chiapas, Quintana Roo, Tabasco, Yucatan) | 6-17 | 3 | 550 | IgG ELISA | [44] | 2016 | C | 0.118 (0.083, 0.181) |
| **Mexico** | Yucatan | 0-50 | 5 | 1,667 | indirect IgG ELISA | [45] | 2014 | A | 0.045 (0.020, 0.081) |
| **Mexico** | State of Morelos | 5-89 | 15 | 928 | indirect IgG ELISA | [46] | 2011 | A | 0.044 (0.018, 0.087) |
| **Pakistan** | Lahore | 0-12 | 3 | 400 | IgG ELISA | [47] | 2010-2015 | A | 0.060 (0.050, 0.070) |
| **Saudi Arabia** | Makkah, Madinah, Jeddah, Jizan | 0-30+ | 3 | 6,397 | capture IgG ELISA | [48] | 2016-2017 | A | 0.008 (0.008, 0.009) |
| **Saudi Arabia** | Jeddah | 0-50+ | 6 | 1,904 | Enzyme immunoassay (EIA) | [49] | 2015* | A | 0.017 (0.016, 0.018) |
| **Singapore** | national | 2-15 | 3 | 1,200 | IgG ELISA | [50] | 2008-2010 | A | 0.020 (0.013, 0.030) |
| **Singapore** | blood service | 16-60 | 9 | 3,627 | IgG ELISA | [51] | 2009-2010 | A | 0.049 (0.012, 0.148) |
| **Singapore** | national | 18-79 | 6 | 1,000 | IgG ELISA | [52] | 2010 | B | 0.034 ( 0.007 , 0.143 ) |
| **Sri Lanka** | City of Colombo | 01-9 | 5 | 799 | IgG ELISA | [53] | 2008-2010 | A | 0.149 (0.076, 0.248) |
| **Taiwan** | Taipei, Taoyuan, Tainan | 0-70+ | 8 | 1,308 | IgG ELISA | [54] | 2010 | A | 0.004 (0.002, 0.011) |
| **Taiwan** | Nanzih district, Kaohsiung City | 0-79 | 7 | 154 | IgG ELISA | [55] | 2015-2016 | A | 0.001 (0.001, 0.001) |
| **Taiwan** | Sanmin district, Kaohsiung City | 0-79 | 7 | 132 | IgG ELISA | [55] | 2015-2016 | A | 0.005 (0.003, 0.007) |
| **Tanzania** | Buhigwe, Kalambo, Kilindi, Kinondoni, Kondoia, Kyela, Mvomero and Ukerewe | 0-42+ | 3 | 1818 | IgG ELISA | [56] | 2018 | A | 0.011 (0.003, 0.035) |
| **Tanzania** | Zanzibar | 0-36+ | 3 | 500 | IgG ELISA | [57] | 2010 | B | 0.114 ( 0.034 , 0.254) |
| **Thailand** | Mukdahn, Ubon, Ratchathani, Savannakhet and Champasak | 5-20+ | 4 | 975 | IgG ELISA | [58] | 2019 | A | 0.083 ( 0.074 , 0.095 ) |
| **Thailand** | Ayutthaya, Lop Buri, Narathiwat and Trag | 0-45+ | 3 | 835 | IgG ELISA | [59] | 2014 | A | 0.070 ( 0.063 , 0.077 ) |
| **Thailand** | Ratchaburi province | 1-55 | 6 | 1,814 | IgG ELISA | [60] | 2012-2015 | A | 0.136 ( 0.127 , 0.146 ) |
| **Thailand** | Ratchaburi province | 9-22 | 3 | 115 | IgG ELISA | [61] | 2019-2020 | A | 0.094 (0.074, 0.118) |
| **Venezuela** | Cana de Azucar | 5-30 | 5 | 2,002 | Hemoagglutinin<br>inibition | [62] | 2010-2011 | **B** | 0.059 ( 0.015 ,<br>0.181 ) |

\*The date of the survey was not specified, instead we report the year of publication of the article.

### FOI estimates

Table 1 and Fig 3 summarise the FOI estimates obtained with the best model selected according to the DIC. The FOI estimates and DIC values obtained with each model version are presented in the Supplementary Figure S1 and Supplementary Tables S3-S4.

**Fig 2.**
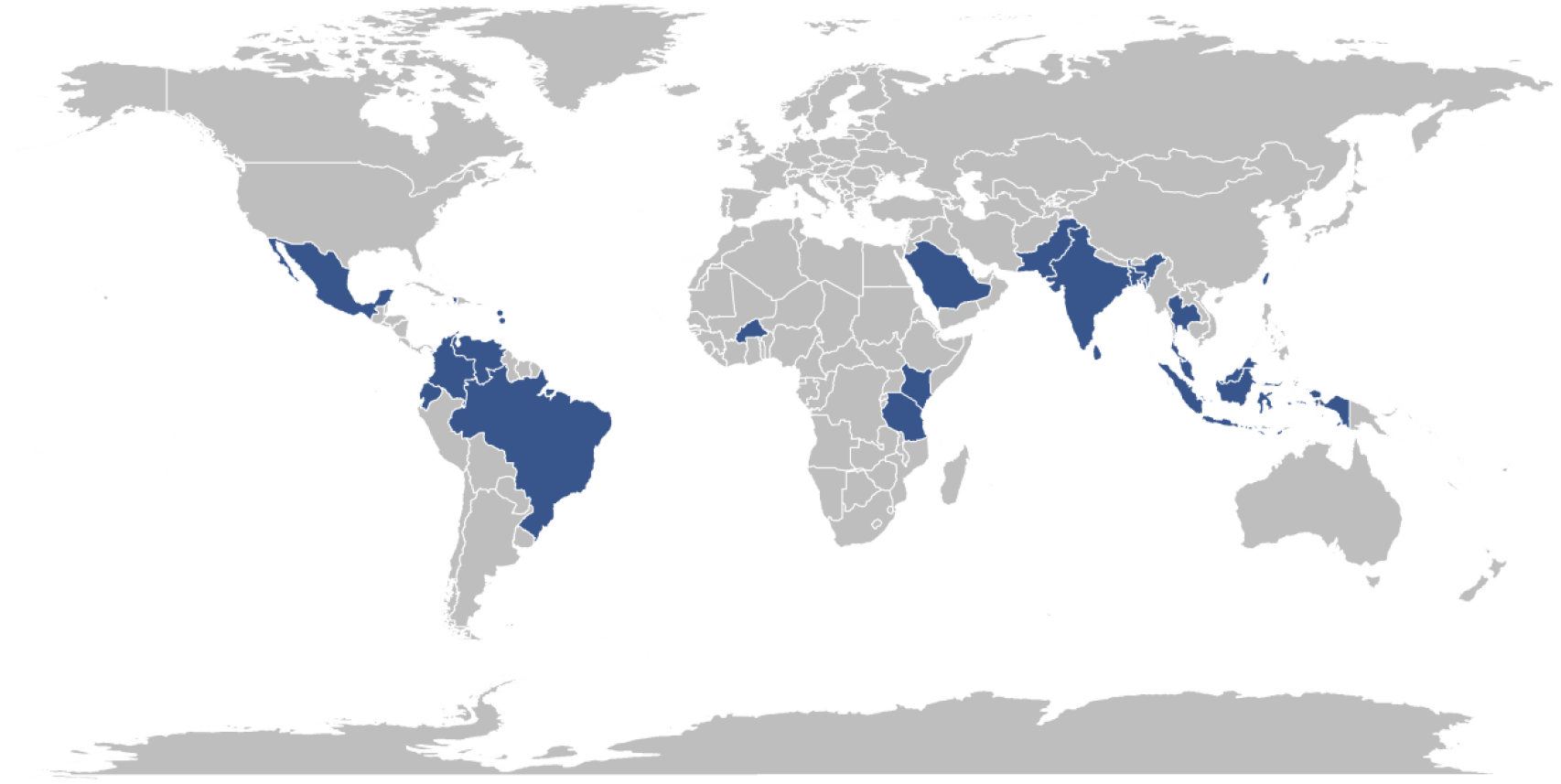
Countries reporting dengue seroprevalence studies from 2014 onwards. World map highlighting in blue the countries with new dengue age-stratified seroprevalence data identified in the literature search.

**Fig 3.**
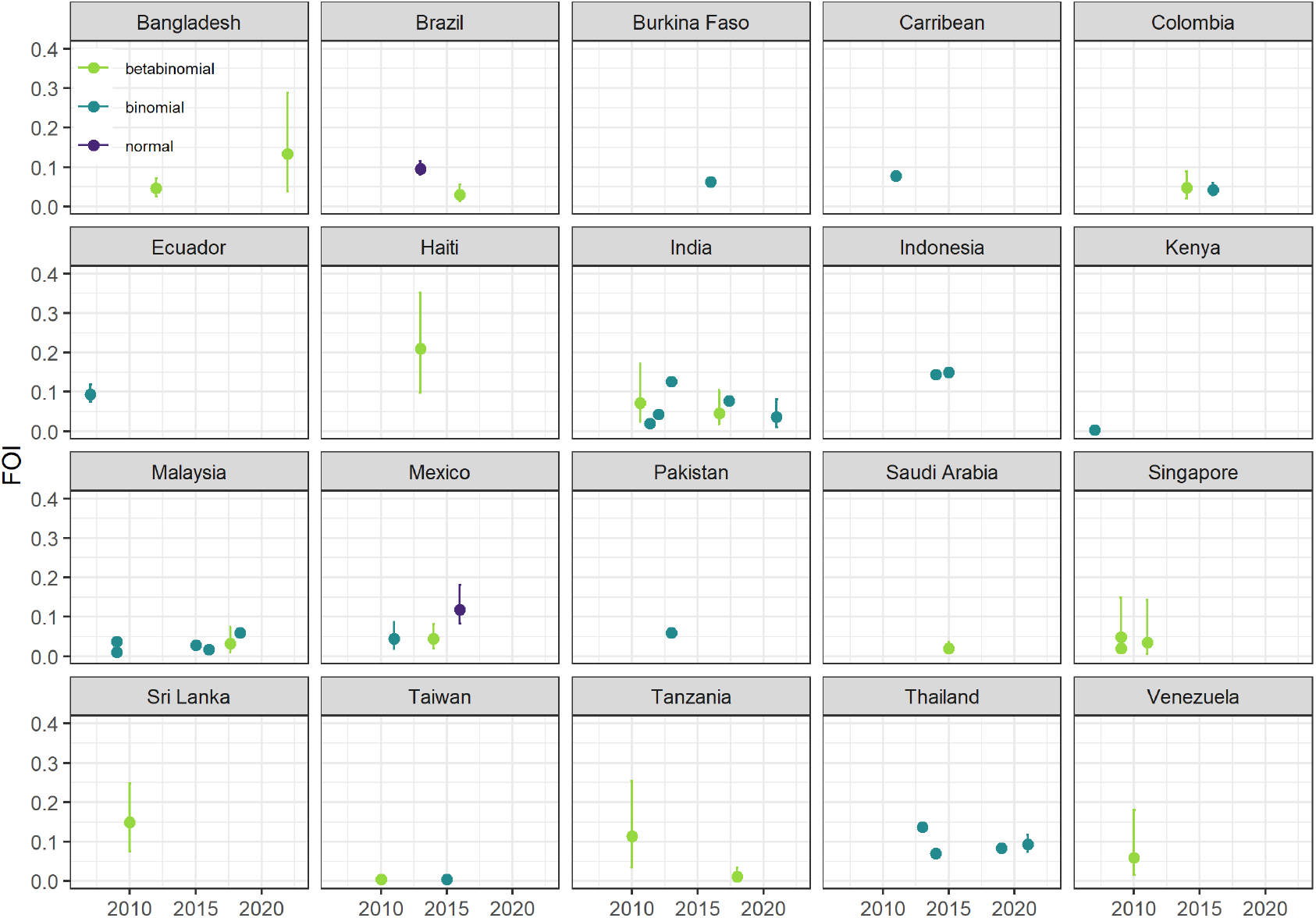
FOI estimates for each dataset by country under the best fitting model. The x axis reports the year of the serological survey. Each panel shows the median (point) and 95% CrI (line) of the FOI estimates by year (x-axis) using a binomial (green), beta-binomial (light blue) and normal (purple) likelihood.

The 44 papers analysed in this study contained a total of 47 dengue seroprevalence datasets and, for the majority of them, the FOI estimates were obtained using model A (Table 1). For the 4 datasets which only reported the positivity rates (not the positive number of subjects among those tested) the FOI was estimated with model C. Overall, Sri Lanka, Thailand, Haiti and Indonesia showed the highest yearly FOI estimates, with values equal to or above 0.14, which correspond to an expected average age of (first) infection of around 7 years. On the other hand, Taiwan was the country with the lowest estimated yearly FOI, around 0.001.

Fig 4 shows the geographical distribution of the dengue serosurveys identified in this analysis and the resulting FOI estimates.

**Fig 4.**
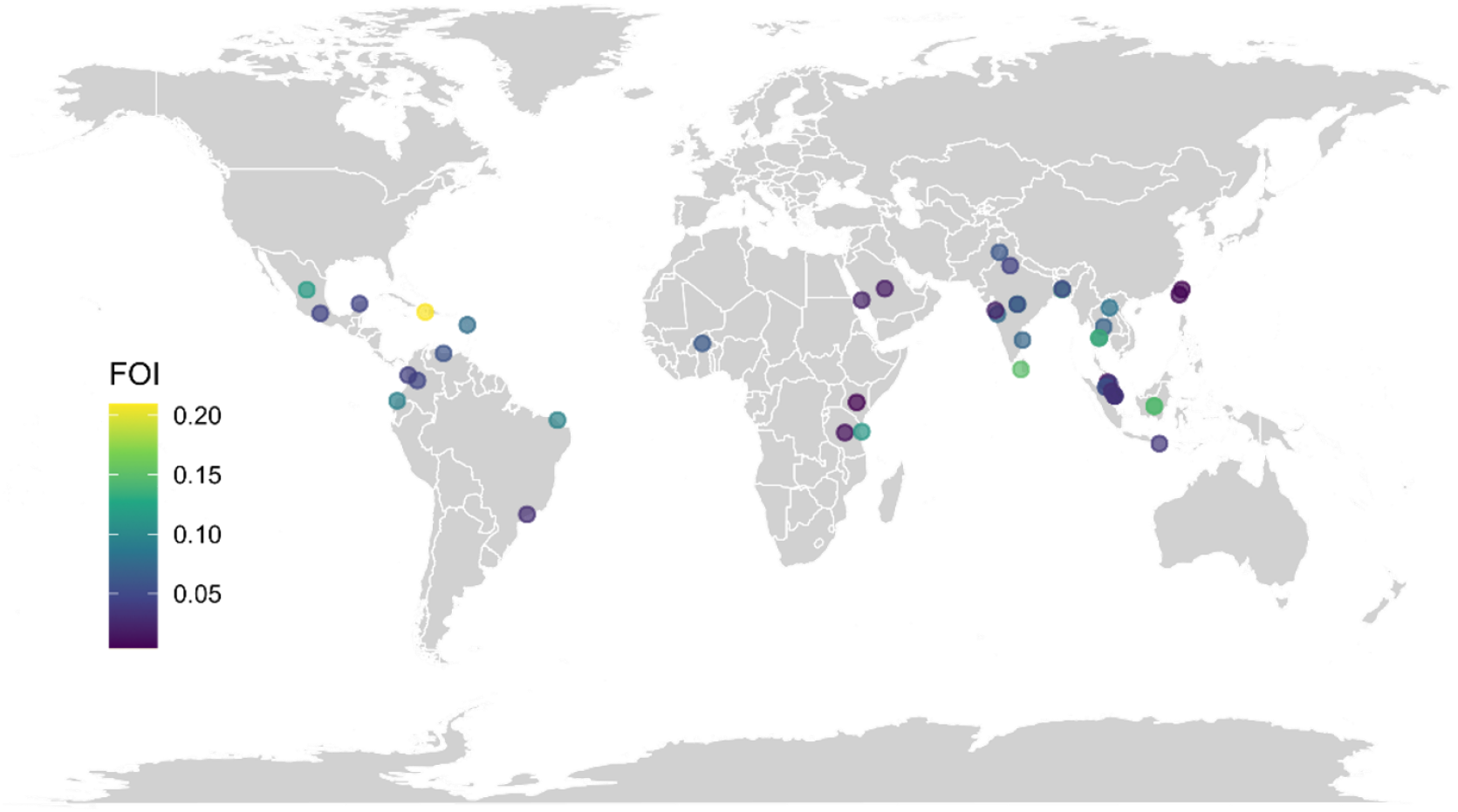
Worldwide distribution and geolocation of the new force of infection (FOI) estimates obtained in this study. Average FOI estimates obtained from the data and models summarized in Table 1.

## Discussion

Age-stratified serological surveys represent an ideal surveillance tool to estimate the risk of dengue infection and reconstruct the age-specific susceptibility profile and the historical circulation of dengue in the surveyed populations [58]. In this study, 44 publications were retrieved from published databases, which provided 47 age-stratified dengue seroprevalence datasets collected between 2007 and 2022 from 20 different countries.

We found that the FOI estimates obtained with models A1 or A2 and B1 or B2 were consistent (Fig S8). As expected, we found high heterogeneities in the average FOI within and between countries [3]. The newly identified age-stratified seroprevalence datasets from the Ratchaburi province of Thailand and at the national level in Singapore generated FOI estimates that were consistent with the estimates obtained from previous studies [4]. On the other hand, the FOI estimates obtained in Lahore (Pakistan) and Colombo city (Sri Lanka) were higher than those estimated in the same territories by Imai et al [4], highlighting within-country heterogeneities in FOI, potentially linked with the observed geographical expansion of dengue circulation [31], and increases in urbanization and population density [35]. As for the other countries previously studied in Imai et al [4] – Brazil, India, Indonesia, Haiti and Mexico – we identified new age-stratified serosurveys, but they were performed in different locations and gave higher FOI estimates.

This study also identified the countries where age-stratified dengue seroprevalence surveys were conducted for the first time since 2014. These include several endemic countries from Central and South America (Ecuador, Guadeloupe, Martinique, Colombia and Venezuela), as well as in Asia (Malaysia, Bangladesh, Taiwan and Saudi Arabia), and Africa (Burkina Faso, Tanzania, Kenya). Of these, the ones from Ecuador, Guadeloupe, Martinique and Burkina Faso are the first to be published in the respective countries [15]. In line with our current understanding of the global trends of dengue transmission, we find evidence of a high risk of infection across Sri Lanka, India, Thailand and Indonesia and lower transmission intensities in countries such as Taiwan (median FOI between 0.001 and 0.005) and Saudi Arabia (median FOI between 0.008 and 0.017), which are located on the outskirts of tropical and subtropical regions (Fig 4). The patterns in Latin America and the Caribbean are more heterogenous, with Haiti showing the highest FOI estimates of this study, with a median FOI estimate of 0.209 (95% CrI 0.1, 0.352).

Through our literature review, we found two age-stratified dengue seroprevalence surveys conducted in Bangladesh, one in Dhaka city conducted in 2012 [20] and one conducted at the national level in 2022 [19]. These datasets report rather different age-dependent susceptibility profiles, with the national survey (performed 10 years after the one in Dhaka city) reporting a higher FOI estimate. Whilst the seroprevalence survey conducted in Dhaka city was a household serosurvey and included all age-groups, the national survey was conducted in blood donors (and did not include individuals <18 years); differences in the surveyed populations could potentially bias the age-stratified seroprevalence profile, but this result suggests an increase in dengue transmission intensity across the country over the last 10 years.

Before this study, we had identified 13 published age-stratified seroprevalence surveys from Africa, specifically from Cameroon, Namibia, Nigeria, Kenya, Sudan and Tanzania [15]. This review identified 4 other age-stratified seroprevalence datasets from the African Region: a national survey in Kenya, two local surveys in Ougadougou (Burkina Faso) and Zanzibar (Tanzania) and a multi-regional survey in Tanzania (Buhigwe, Kalambo, Kilindi, Kinondoni, Kondoa, Kyela, Mvomero and Ukerewe). Amongst all the FOI estimates obtained across Africa, the serosurvey conducted in 2010 in Tanzania shows the highest FOI estimate (median 0.114 (95% CrI 0.034, 0.254)), consistent with estimates obtained in blood donors [57].

Notably, we found two other age-stratified dengue seroprevalence surveys from Tanzania, one at a multi-regional level [56] and one in Zanzibar [57], reporting rather different age-dependent susceptibility profiles. The multi-regional survey performed in 2018 suggests a relatively recent introduction of dengue (flat immunity profile), whilst the results reported in Zanzibar in 2010 show clear age-dependencies in the seroprevalence. These differences could reflect fundamental differences in the transmission intensity, given that *Aedes aegypti* mosquitoes are urban vectors, and hence dengue is expected to circulate at higher intensity in urban settings, whilst the multi-regional seroprevalence profile likely captures the average immunity profile of the population at the national level, including in rural settings where dengue is likely to be circulating at lower intensities.

The dengue FOI estimates obtained in this study, along with the estimates obtained in previous analyses [15] (Table S9), provide a compendium of the global FOI estimates obtained from age-stratified seroprevalence surveys using catalytic models, which will be useful for guiding future intervention and vaccination strategies.

## Conclusion

Age-stratified serological surveys provide key epidemiological data for estimating the transmission intensity of dengue, which in turn can be used to refine burden estimates and inform the optimal implementation of interventions such as vaccination. Our findings, alongside the serological data and FOI estimates published in Imai et al [4] and Cattarino et al [15], summarize our current understanding of global heterogeneities in transmission intensity, as well as disparities in the geographical distribution of age-stratified serological surveys, thus highlighting regional gaps in arbovirus surveillance, which can help inform current and future efforts to strengthen surveillance capacity locally and globally.

## Data Availability

All data produced in the present work are contained in the manuscript.

## Author contribution

Data curation: AV, ID

Formal analysis: AV, ID

Methodology: AV, ID

Software: AV, ID

Validation: AV, CMC

Visualization: AV, CMC, ID

Writing- Original Draft Preparation: AV, CMC, ID

Writing. review & editing: AV, CMC, ID, BP, IR, NM

Funding acquisition: ID, BP, IR, NM

Supervision: ID

Conceptualization: ID, BP, IR, NM

