## Supplementary Information for "A scoping literature review of global dengue age-stratified seroprevalence data: estimating dengue force of infection in endemic countries"

#### Table of Contents

Tables…………………………………………………………………………………………...2
Figures………………………………………………………………………………………16

### Tables

The literature search was performed on the 29th of September 2022, as reported in Tables S1 and S2.

**Table S1. Literature search performed in Embase.**

| Step | Search Statement |
|---|---|
| 1 | exp Dengue virus/ or exp dengue/ or exp severe dengue/ or dengue.mp. |
| 2 | seropositiv*.mp. |
| 3 | positiv*.mp. |
| 4 | seroprevalence.mp. or exp seroprevalence/ |
| 5 | sero*.mp. |
| 6 | 2 or 3 or 4 |
| 7 | 1 and 5 and 6 |
| 8 | limit 7 to yr=”2014 -Current” |

**Table S2. Literature search performed in Medline.**

| Step | Search Statement |
|---|---|
| 1 | dengue.mp. or exp Dengue/ or Severe Dengue/ or Dengue Virus/ |
| 2 | sero*.mp. |
| 3 | Prevalence/ or prevalence.mp. |
| 4 | seroprevalence.mp. or Seroepidemiologic Studies/ |
| 5 | positiv*.mp. |
| 6 | seropositiv*.mp. |
| 7 | 3 or 4 or 5 or 6 |
| 8 | 1 and 2 and 7 |
| 9 | limit 8 to yr=”2014 -Current” |

**Table S3. Region-specific FOI estimates obtained under model A.** Specifically, the table reports the FOI estimated under model A1 (binomial likelihood) and model A2 (betabinomial likelihood) with constant FOI. The DIC (deviance information criterion) of the two modelsis also reported to compare them.

| Country | Location | FOI (model A1) | FOI (model A2) | DIC (model A1) | DIC (model A2) | Ref. |
|---|---|---|---|---|---|---|
| **Bangladesh** | Dhaka city | 0.064 ( 0.059 , 0.069 ) | 0.045 (0.026, 0.071) | 100 | 61 | [20] |
| **Brazil** | Sau Paulo | 0.035 ( 0.033 , 0.038 ) | 0.030 (0.013, 0.055) | 116 | 48 | [21] |
| **Burkina Faso** | Ougadougou | 0.063 ( 0.059 , 0.066 ) | 0.054 (0.035, 0.076) | 54 | 83 | [23] |
| **Colombia** | Quobdo | 0.041 ( 0.028 , 0.06 ) | 0.062 (0.024, 0.139) | 15 | 21 | [25] |
| **Colombia** | Anapoima, Apulo, Buenaventura, Quibdo, Tumaco, Tierralta | 0.099 ( 0.091 , 0.107 ) | 0.047 (0.020, 0.088) | 333 | 50 | [24] |
| **Ecuador** | Town of Quininde | 0.094 ( 0.074 , 0.118 ) | 0.084 (0.035, 0.159) | 18 | 23 | [26] |
| **France (Carribean)** | Guadeloupe and Martinique | 0.078 ( 0.07 , 0.087 ) | 0.048 (0.026, 0.073) | 40 | 47 | [27] |
| **India** | Punjab, NCT of Delhi, Uttar Pradesh, Tripura, Meghalaya, Assam, Bihar, West Bengal, Odisha, Rajasthan, Madhya Pradesh, Maharashtra, Andhra Pradesh, Karnataka, Tamil Nadu | 0.034 ( 0.033 , 0.035 ) | 0.046 (0.017, 0.105) | 400 | 53 | [30] |
| **India** | Denpasar Bali | 0.061 ( 0.055 , 0.069 ) | 0.036 (0.011, 0.081) | 80 | 25 | [33] |
| **India** | New Delhi, Hyderabad, Kalyani, Wardha, mumbai, Bangalore | 0.127 ( 0.121 , 0.134 ) | 0.121 (0.071, 0.187) | 53 | 71 | [29] |
| **India** | Delhi | 0.043 ( 0.036 , 0.052 ) | 0.045 (0.020, 0.087) | 16 | 25 | [32] |
| **India** | Vadu area | 0.021 ( 0.018 , 0.023 ) | 0.024 (0.012, 0.047) | 47 | 50 | [35] |
| **India** | Pune | 0.078 ( 0.072 , 0.084 ) | 0.063 (0.045, 0.084) | 63 | 91 | [34] |
| **Indonesia** | national | 0.144 ( 0.138 , 0.151 ) | 0.122 (0.062, 0.202) | 34 | 53 | [36] |
| **Indonesia** | national | 0.149 ( 0.142 , 0.156 ) | 0.138 (0.111, 0.168) | 129 | 154 | [37] |
| **Kenya** | national | 0.004 ( 0.004 , 0.005 ) | 0.011 (0.003, 0.033) | 19 | 35 | [41] |
| **Malaysia** | Peninsular Malaysia | 0.038 ( 0.036 , 0.041 ) | 0.029 (0.014, 0.049) | 36 | 42 | [41] |
| **Malaysia** | Petaling district | 0.059 ( 0.052 , 0.066 ) | 0.043 (0.025, 0.066) | 88 | 65 | [42] |
| **Malaysia** | Federal territory of Kuala, Lumpur, Perak, Kedah, Penang, Johor, Pahang, Kelantan and Sabah | 0.010 ( 0.008 , 0.012 ) | 0.027 (0.009, 0.078) | 31 | 45 | [39] |
| **Malaysia** | Damansara Damai | 0.042 ( 0.031 , 0.055 ) | 0.033 (0.010, 0.075) | 21 | 19 | [40] |
| **Malaysia** | Sungai Segamat | 0.029 ( 0.025 , 0.033 ) | 0.027 (0.014, 0.046) | 37 | 39 | [43] |

| | | | | | | |
|---|---|---|---|---|---|---|
| **Mexico** | state of Morelos | 0.052 ( 0.047 , 0.057 ) | 0.044 (0.018, 0.087) | 229 | 48 | [46] |
| **Mexico** | Yucatan | 0.056 ( 0.053 , 0.06 ) | 0.045 (0.020, 0.081) | 362 | 64 | [45] |
| **Pakistan** | Lahore | 0.060 (0.050, 0.070) | 0.088 (0.033, 0.193) | 23 | 34 | [47] |
| **Saudi Arabia** | Jeddah | 0.017 (0.016, 0.018) | 0.020 (0.011, 0.037) | 26 | 32 | [49] |
| **Saudi Arabia** | Makkah, Madinah, Jeddah, Jizan | 0.008 (0.008, 0.009) | 0.020 (0.006, 0.057) | 56 | 65 | [48] |
| **Singapore** | national | 0.020 (0.019, 0.021) | 0.020 (0.013, 0.030) | 299 | 102 | [50] |
| **Singapore** | blood service | 0.013 (0.011, 0.016) | 0.049 (0.012, 0.148) | 109 | 37 | [51] |
| **Sri Lanka** | city of Colombo | 0.155 (0.140, 0.171) | 0.149 (0.076, 0.248) | 55 | 51 | [53] |
| **Taiwan** | Nanzih district,Kaohsiung City | 0.001 (0.001, 0.001) | 0.004 (0.002, 0.011) | 31 | 37 | [55] |
| **Taiwan** | Sanmin district,Kaohsiung City | 0.005 (0.003, 0.007) | 0.009 (0.004, 0.019) | 20 | 32 | [56] |
| **Taiwan** | Taipei, Taoyuan, Tainan | 0.002 (0.001, 0.004) | 0.004 (0.002, 0.011) | 63 | 61 | [54] |
| **Tanzania** | Buhigwe, Kalambo,Kilindi, Kinondoni, Kondoa, Kyela, Mvomero and Ukerewe | 0.005 (0.004, 0.005) | 0.011 (0.003, 0.035) | 70 | 40 | [56] |
| **Thailand** | Ratchaburi province | 0.094 (0.074, 0.118) | 0.083 (0.035, 0.156) | 18 | 23 | [60] |
| **Thailand** | Ratchaburi province | 0.136 ( 0.127 , 0.146 ) | 0.089 (0.050, 0.134) | 53 | 60 | [62] |
| **Thailand** | Ayutthaya, Lop Buri, | 0.07 ( 0.063 , 0.077 ) | 0.045 ( 0.02 , 0.085 ) | 18 | 31 | [59] |
| **Thailand** | Mukdahn, Ubon Ratchathani, Savannakhet and Champasak | 0.083 ( 0.074 , 0.095 ) | 0.061 ( 0.03 , 0.104 ) | 21 | 36 | [58] |

**Table S4. Region-specific FOI estimates obtained under model B.** Specifically, the table reports the FOI estimated under model B1 (binomial likelihood) and model B2 (betabinomial likelihood) with constant FOI and antibody decay. The DIC (deviance information criterion) of the two models is also reported to compare them.

| Country | Location | FOI (model B1) | FOI (model B2) | DIC (model B1) | DIC (model B2) | Ref. |
|---|---|---|---|---|---|---|
| **Bangladesh** | blood service | 0.111 ( 0.053 , 0.19 ) | 0.133 ( 0.038 , 0.288) ) | 1498 | 79 | [19] |
| **Haiti** | Gressier, Jacmel, Chabin | 0.292 ( 0.213 , 0.414 ) | 0.209 ( 0.1 , 0.351 ) | 1175 | 128 | [28] |
| **India** | Chennai | 0.067 ( 0.042 , 0.101 ) | 0.072 ( 0.024 , 0.172 ) | 1911 | 123 | [31] |
| **Singapore** | national | 0.005 ( 0.002 , 0.01 ) | 0.034 ( 0.007 , 0.143 ) | 360 | 81 | [52] |
| **Tanzania** | Zanzibar | 0.109 ( 0.066 , 0.24 ) | 0.114 ( 0.034 , 0.254 ) | 1613 | 59 | [57] |
| **Venezuela** | Cana de Azucar | 0.020 ( 0.012 , 0.034 ) | 0.059 ( 0.015 , 0.181 ) | 576 | 70 | [62] |

**Table S5. Region-specific FOI estimates obtained under model A1.** Specifically, the table reports the FOI estimated under model A1 (binomial likelihood) together with -the age at which we expect 50% seroprevalence and 70% seroprevalence and the average age at first infections. “NA” is used when the average age was above 100 years old.

| Country | Region | FOI | age_50%serop | age_70%serop | age_first_infection |
|---|---|---|---|---|---|
| **Bangladesh** | Dhaka city | 0.064 ( 0.059 , 0.069 ) | 11 ( 10 , 12 ) | 19 ( 18 , 20 ) | 16 ( 15 , 17 ) |
| **Brazil** | Sau Paulo | 0.035 ( 0.033 , 0.038 ) | 20 ( 18 , 21 ) | 34 ( 31 , 36 ) | 28 ( 26 , 30 ) |
| **Burkina Faso** | Ougadougou | 0.063 ( 0.059 , 0.066 ) | 11 ( 11 , 12 ) | 19 ( 18 , 20 ) | 16 ( 15 , 17 ) |
| **Colombia** | Anapoima, Apulo, Buenaventura, Quibdo, Tumaco, Tierralta | 0.099 ( 0.091 , 0.107 ) | 7 ( 7 , 7 ) | 12 ( 11 , 13 ) | 10 ( 9 , 11 ) |
| **Colombia** | Quobdo | 0.041 ( 0.028 , 0.06 ) | 17 ( 11 , 25 ) | 29 ( 20 , 44 ) | 24 ( 16 , 36 ) |
| **Ecuador** | Town of Quininde | 0.094 ( 0.074 , 0.118 ) | 7 ( 6 , 9 ) | 13 ( 10 , 16 ) | 11 ( 8 , 13 ) |
| **French Carribean** | Guadeloupe and Martinique | 0.078 ( 0.07 , 0.087 ) | 9 ( 8 , 10 ) | 15 ( 14 , 17 ) | 13 ( 12 , 14 ) |
| **India** | New Delhi, Hyderabad, Kalyani, Wardha, mumbai, Bangalore | 0.127 ( 0.121 , 0.134 ) | 5 ( 5 , 6 ) | 9 ( 9 , 10 ) | 8 ( 7 , 8 ) |
| **India** | Punjab, NCT of Delhi, Uttar Pradesh, Tripura, Meghalaya, Assam, Bihar, West Bengal, Odisha, Rajasthan, Madhya Pradesh, Maharashtra, Andhra Pradesh, Karnataka, Tamil Nadu | 0.034 ( 0.033 , 0.035 ) | 20 ( 20 , 21 ) | 35 ( 34 , 36 ) | 29 ( 28 , 30 ) |
| **India** | Delhi | 0.043 ( 0.036 , 0.052 ) | 16 ( 14 , 20 ) | 28 ( 24 , 34 ) | 23 ( 20 , 28 ) |
| **India** | Denpasar Bali | 0.061 ( 0.055 , 0.069 ) | 11 ( 10 , 13 ) | 20 ( 18 , 22 ) | 17 ( 15 , 18 ) |

| | | | | | |
|---|---|---|---|---|---|
| **India** | Pune | 0.078 ( 0.072 , 0.084 ) | 9 ( 8 , 10 ) | 16 ( 14 , 17 ) | 13 ( 12 , 14 ) |
| **India** | Vadu area | 0.021 ( 0.018 , 0.023 ) | 34 ( 30 , 37 ) | 59 ( 53 , 64 ) | 49 ( 44 , 53 ) |
| **Indonesia** | national | 0.149 ( 0.142 , 0.156 ) | 5 ( 4 , 5 ) | 8 ( 8 , 8 ) | 7 ( 6 , 7 ) |
| **Indonesia** | national | 0.144 ( 0.138 , 0.151 ) | 5 ( 5 , 5 ) | 8 ( 8 , 9 ) | 7 ( 7 , 7 ) |
| **Kenya** | national | 0.004 ( 0.004 , 0.005 ) | NA | NA | NA |
| **Malaysia** | Federal territory of Kuala Lumpur, Perak, Kedah, Penang, Johor, Pahang, Kelantan and Sabah | 0.01 ( 0.008 , 0.012 ) | 70 ( 61 , 80 ) | NA | NA |
| **Malaysia** | Damansara Damai | 0.042 ( 0.031 , 0.055 ) | 17 ( 13 , 21 ) | 29 ( 23 , 37 ) | 24 ( 19 , 31 ) |
| **Malaysia** | Peninsular Malaysia | 0.038 ( 0.036 , 0.041 ) | 18 ( 17 , 19 ) | 31 ( 30 , 34 ) | 26 ( 25 , 28 ) |
| **Malaysia** | Petaling district | 0.059 ( 0.052 , 0.066 ) | 12 ( 11 , 13 ) | 21 ( 18 , 22 ) | 17 ( 15 , 19 ) |
| **Malaysia** | Sungai Segamat | 0.029 ( 0.025 , 0.033 ) | 24 ( 21 , 29 ) | 42 ( 36 , 51 ) | 35 ( 30 , 42 ) |
| **Mexico** | Yucatan | 0.056 ( 0.053 , 0.06 ) | 12 ( 12 , 13 ) | 21 ( 20 , 23 ) | 18 ( 17 , 19 ) |
| **Mexico** | state of Morelos | 0.052 ( 0.047 , 0.057 ) | 13 ( 12 , 14 ) | 23 ( 22 , 25 ) | 19 ( 18 , 21 ) |
| **Pakistan** | Lahore | 0.06 ( 0.05 , 0.07 ) | 12 ( 10 , 13 ) | 20 ( 17 , 23 ) | 17 ( 14 , 19 ) |
| **Saudi Arabia** | Makkah, Madinah, Jeddah, Jizan | 0.008 ( 0.008 , 0.009 ) | 85 ( 82 , 89 ) | NA | NA |
| **Saudi Arabia** | Jeddah | 0.017 ( 0.016 , 0.018 ) | 41 ( 38 , 43 ) | 71 ( 66 , 75 ) | 59 ( 55 , 63 ) |
| **Singapore** | blood service | 0.013 ( 0.011 , 0.016 ) | 52 ( 45 , 61 ) | 89 ( 78 , 106 ) | 74 ( 65 , 88 ) |
| **Singapore** | national | 0.02 ( 0.019 , 0.021 ) | 34 ( 33 , 36 ) | 59 ( 57 , 63 ) | 49 ( 47 , 52 ) |
| **Sri Lanka** | city of Colombo | 0.155 ( 0.14 , 0.171 ) | 4 ( 4 , 5 ) | 8 ( 7 , 9 ) | 6 ( 6 , 7 ) |
| **Taiwan** | Taipei, Taoyuan, Tainan | 0.001 ( 0.001 , 0.001 ) | NA | NA | NA |
| **Taiwan** | Nanzih district, Kaohsiung City | 0.005 ( 0.003 , 0.007 ) | NA | NA | NA |
| **Taiwan** | Sanmin district, Kaohsiung City | 0.002 ( 0.001 , 0.004 ) | NA | NA | NA |
| **Tanzania** | Buhigwe, Kalambo, Kilindi, Kinondoni, Kondoa, Kyela, Mvomero and Ukerewen | 0.005 ( 0.004 , 0.005 ) | NA | NA | NA |
| **Thailand** | Ratchaburi province | 0.094 ( 0.074 , 0.118 ) | 7 ( 6 , 9 ) | 13 ( 10 , 16 ) | 11 ( 8 , 13 ) |
| **Thailand** | Ayutthaya, Lop Buri, Narathiwat and Trag | 0.07 ( 0.063 , 0.077 ) | 10 ( 9 , 11 ) | 17 ( 16 , 19 ) | 14 ( 13 , 16 ) |
| **Thailand** | Mukdahn, Ubon Ratchathani, Savannakhet and Champasak | 0.083 ( 0.074 , 0.095 ) | 8 ( 7 , 9 ) | 14 ( 12 , 16 ) | 12 ( 10 , 13 ) |
| **Thailand** | Ratchaburi province | 0.136 ( 0.127 , 0.146 ) | 5 ( 5 , 5 ) | 9 ( 8 , 9 ) | 7 ( 7 , 8 ) |

**Table S6. Region-specific FOI estimates obtained under model A2.** Specifically, the table reports the FOI estimated under model A2 (betabinomial likelihood) together with -the age at which we expect 50% seroprevalence and 70% seroprevalence and the average age at first infections. The table also reports the estimates of the overdispersion parameter phi obtained under model A2. “NA” is used when the average age was above 100 years old.

| Country | Region | FOI | age_50%serop | age_70%serop | age_first_infection | phi |
|---|---|---|---|---|---|---|
| **Bangladesh** | Dhaka city | 0.045 ( 0.026 , 0.071 ) | 16 ( 10 , 26 ) | 27 ( 17 , 44 ) | 22 ( 14 , 37 ) | 0.243 ( 0.123 , 0.483 ) |
| **Brazil** | Sau Paulo | 0.03 ( 0.013 , 0.055 ) | 21 ( 12 , 49 ) | 37 ( 20 , 85 ) | 31 ( 17 , 70 ) | 0.285 ( 0.136 , 0.571 ) |
| **Burkina Faso** | Ougadougou | 0.054 ( 0.035 , 0.076 ) | 13 ( 9 , 19 ) | 22 ( 16 , 33 ) | 18 ( 13 , 27 ) | 0.17 ( 0.089 , 0.342 ) |
| **Colombia** | Anapoima, Apulo, Buenaventura, Quibdo, Tumaco, Tierralta | 0.047 ( 0.02 , 0.088 ) | 14 ( 8 , 33 ) | 24 ( 14 , 57 ) | 20 ( 11 , 48 ) | 0.381 ( 0.185 , 0.681 ) |
| **Colombia** | Quobdo | 0.062 ( 0.024 , 0.139 ) | 12 ( 5 , 26 ) | 21 ( 9 , 45 ) | 18 ( 8 , 37 ) | 0.315 ( 0.147 , 0.635 ) |
| **Ecuador** | Town of Quininde | 0.084 ( 0.035 , 0.159 ) | 8 ( 5 , 17 ) | 14 ( 8 , 29 ) | 12 ( 7 , 24 ) | 0.313 ( 0.141 , 0.637 ) |
| **French Carribean** | Guadeloupe and Martinique | 0.048 ( 0.026 , 0.073 ) | 14 ( 9 , 23 ) | 25 ( 15 , 40 ) | 20 ( 13 , 33 ) | 0.261 ( 0.124 , 0.544 ) |
| **India** | New Delhi, Hyderabad, Kalyani, Wardha, mumbai, Bangalore | 0.121 ( 0.071 , 0.187 ) | 6 ( 3 , 10 ) | 10 ( 6 , 18 ) | 8 ( 5 , 15 ) | 0.212 ( 0.105 , 0.429 ) |
| **India** | Punjab, NCT of Delhi, Uttar Pradesh, Tripura, Meghalaya, Assam, Bihar, West Bengal, Odisha, Rajasthan, Madhya Pradesh, Maharashtra, Andhra Pradesh, Karnataka, Tamil Nadu | 0.046 ( 0.017 , 0.105 ) | 15 ( 7 , 41 ) | 27 ( 12 , 72 ) | 22 ( 10 , 59 ) | 0.309 ( 0.144 , 0.62 ) |
| **India** | Delhi | 0.045 ( 0.02 , 0.087 ) | 16 ( 8 , 31 ) | 28 ( 14 , 53 ) | 23 ( 12 , 44 ) | 0.311 ( 0.143 , 0.641 ) |
| **India** | Denpasar Bali | 0.036 ( 0.011 , 0.081 ) | 20 ( 9 , 53 ) | 35 ( 15 , 92 ) | 29 ( 13 , 76 ) | 0.386 ( 0.166 , 0.745 ) |
| **India** | Pune | 0.063 ( 0.045 , 0.084 ) | 11 ( 8 , 16 ) | 19 ( 15 , 27 ) | 16 ( 12 , 23 ) | 0.142 ( 0.078 , 0.271 ) |
| **India** | Vadu area | 0.024 ( 0.012 , 0.047 ) | 29 ( 18 , 54 ) | 51 ( 31 , 94 ) | 42 ( 26 , 78 ) | 0.238 ( 0.118 , 0.476 ) |
| **Indonesia** | national | 0.138 ( 0.111 , 0.168 ) | 5 ( 4 , 6 ) | 9 ( 7 , 11 ) | 7 ( 6 , 9 ) | 0.103 ( 0.062 , 0.185 ) |
| **Indonesia** | national | 0.122 ( 0.062 , 0.202 ) | 6 ( 4 , 12 ) | 10 ( 6 , 20 ) | 8 ( 5 , 17 ) | 0.255 ( 0.121 , 0.535 ) |
| **Kenya** | national | 0.011 ( 0.003 , 0.033 ) | 64 ( 26 , 179 ) | NA | 92 ( 37 , 259 ) | 0.353 ( 0.151 , 0.691 ) |
| **Malaysia** | Federal territory of Kuala Lumpur, Perak, Kedah, Penang, Johor, Pahang, Kelantan and Sabah | 0.027 ( 0.009 , 0.078 ) | 27 ( 8 , 74 ) | 48 ( 14 , 129 ) | 40 ( 12 , 107 ) | 0.321 ( 0.141 , 0.644 ) |
| **Malaysia** | Damansara Damai | 0.033 ( 0.01 , 0.075 ) | 21 ( 10 , 71 ) | 36 ( 17 , 124) | 30 ( 14 , 103 ) | 0.371 ( 0.162 , 0.737 ) |
| **Malaysia** | Peninsular Malaysia | 0.029 ( 0.014 , 0.049 ) | 22 ( 13 , 47 ) | 39 ( 23 , 81 ) | 32 ( 19 , 67 ) | 0.28 ( 0.128 , 0.582 ) |
| **Malaysia** | Petaling district | 0.043 ( 0.025 , 0.066 ) | 17 ( 10 , 26 ) | 29 ( 18 , 45 ) | 24 ( 15 , 37 ) | 0.246 ( 0.129 , 0.452 ) |

| | | | | | | |
|---|---|---|---|---|---|---|
| **Malaysia** | Sungai Segamat | 0.027 ( 0.014 , 0.046 ) | 26 ( 16 , 48 ) | 45 ( 29 , 83 ) | 37 ( 24 , 69 ) | 0.257 ( 0.123 , 0.516 ) |
| **Mexico** | Yucatan | 0.045 ( 0.02 , 0.081 ) | 16 ( 9 , 29 ) | 28 ( 16 , 51 ) | 23 ( 14 , 42 ) | 0.296 ( 0.15 , 0.557 ) |
| **Mexico** | state of Morelos | 0.044 ( 0.018 , 0.087 ) | 16 ( 8 , 36 ) | 28 ( 14 , 63 ) | 23 ( 12 , 53 ) | 0.293 ( 0.133 , 0.62 ) |
| **Pakistan** | Lahore | 0.088 ( 0.033 , 0.193 ) | 8 ( 4 , 24 ) | 14 ( 7 , 41 ) | 12 ( 6 , 34 ) | 0.316 ( 0.142 , 0.637 ) |
| **Saudi Arabia** | Makkah, Madinah, Jeddah, Jizan | 0.02 ( 0.006 , 0.057 ) | 33 ( 12 , NA ) | 58 ( 20 , 215 ) | 48 ( 17 , 179 ) | 0.34 ( 0.152 , 0.671 ) |
| **Saudi Arabia** | Jeddah | 0.02 ( 0.011 , 0.037 ) | 36 ( 20 , 64 ) | 62 ( 35 , NA ) | 51 ( 29 , 92 ) | 0.221 ( 0.11 , 0.442 ) |
| **Singapore** | blood service | 0.049 ( 0.012 , 0.148 ) | 15 ( 5 , 54 ) | 26 ( 8 , 94 ) | 21 ( 7 , 78 ) | 0.405 ( 0.175 , 0.736 ) |
| **Singapore** | national | 0.02 ( 0.013 , 0.03 ) | 34 ( 24 , 48 ) | 59 ( 42 , 83 ) | 49 ( 35 , 69 ) | 0.202 ( 0.111 , 0.371 ) |
| **Sri Lanka** | city of Colombo | 0.149 ( 0.076 , 0.248 ) | 4 ( 3 , 9 ) | 8 ( 5 , 15 ) | 6 ( 4 , 13 ) | 0.238 ( 0.118 , 0.485 ) |
| **Taiwan** | Taipei, Taoyuan, Tainan | 0.004 ( 0.002 , 0.011 ) | NA | NA | NA | 0.258 ( 0.12 , 0.53 ) |
| **Taiwan** | Nanzih district, Kaohsiung City | 0.009 ( 0.004 , 0.019 ) | 71 ( 39 , 166 ) | NA | NA | 0.283 ( 0.139 , 0.536 ) |
| **Taiwan** | Sanmin district, Kaohsiung City | 0.005 ( 0.002 , 0.012 ) | NA | NA | NA | 0.259 ( 0.123 , 0.521 ) |
| **Tanzania** | Buhigwe, Kalambo, Kilindi, Kinondoni, Kondoa, Kyela, Mvomero and Ukerewen | 0.011 ( 0.003 , 0.035 ) | NA | NA | 87 ( 36 , 390 ) | 0.352 ( 0.154 , 0.698 ) |
| **Thailand** | Ratchaburi province | 0.083 ( 0.035 , 0.156 ) | 8 ( 4 , 23 ) | 14 ( 8 , 40 ) | 12 ( 6 , 33 ) | 0.313 ( 0.144 , 0.642 ) |
| **Thailand** | Ayutthaya, Lop Buri, Narathiwat and Trag | 0.045 ( 0.02 , 0.085 ) | 16 ( 9 , 32 ) | 27 ( 15 , 55 ) | 22 ( 13 , 45 ) | 0.305 ( 0.136 , 0.646 ) |
| **Thailand** | Mukdahn, Ubon Ratchathani, Savannakhet and Champasak | 0.061 ( 0.03 , 0.104 ) | 12 ( 7 , 20 ) | 21 ( 11 , 35 ) | 17 ( 10 , 29 ) | 0.278 ( 0.128 , 0.573 ) |
| **Thailand** | Ratchaburi province | 0.089 ( 0.05 , 0.134 ) | 8 ( 5 , 13 ) | 14 ( 9 , 22 ) | 12 ( 8 , 18 ) | 0.239 ( 0.115 , 0.496 ) |

**Table S7. Region-specific FOI estimates obtained under model B1.** Specifically, the table reports the FOI estimated under model A1 (binomial likelihood) together with -the age at which we expect 50% seroprevalence and 70% seroprevalence and the average age at first infections. The table also reports the estimates of the age decay alpha parameter obtained under model B1. "NA" is used when the average age was above 100 years old.

| Country | Location | FOI | age_50%serop | age_70%serop | age_first_infection | alpha |
|---|---|---|---|---|---|---|
| **Bangladesh** | blood service | 0.111 ( 0.053 , 0.19 ) | NA | NA | 9 ( 5 , 19 ) | 0.2 ( 0.09 , 0.34 ) |
| **Haiti** | Gressier, Jacmel, Chabin | 0.292 ( 0.213 , 0.414 ) | 3 ( 2, 4 ) | 8 ( 6 , 12 ) | 4 ( 3 , 5 ) | 0.11 ( 0.07 , 0.16 ) |
| **India** | Chennai | 0.067 ( 0.042 , 0.101 ) | NA | NA | 14 ( 11 , 24 ) | 0.28 ( 0.18 , 0.42 ) |
| **Singapore** | national | 0.005 ( 0.002 , 0.01 ) | 0 | NA | NA | 0.19 ( 0.08 , 0.35 ) |
| **Tanzania** | Zanzibar | 0.109 ( 0.066 , 0.24 ) | NA | NA | 9 ( 14 , 16 ) | 0.1 ( 0.05 , 0.23 ) |
| **Venezuela** | Cana de Azucar | 0.02 ( 0.012 , 0.034 ) | NA | NA | 52 ( 31 , 74 ) | 0.16 ( 0.08 , 0.3 ) |

**Table S8. Region-specific FOI estimates obtained under model B2.** Specifically, the table reports the FOI estimated under model A1 (betabinomial likelihood) together with -the age at which we expect 50% seroprevalence and 70% seroprevalence and the average age at first infections. The table also reports the estimates of the age decay alpha parameter and the overdispersion parameter phi obtained under model B2. "NA" is used when the average age was above 100 years old.

| Country | Location | FOI | age_50% serop | age_70% serop | age_first_ infection | alpha | phi |
|---|---|---|---|---|---|---|---|
| **Bangladesh** | blood service | 0.133 ( 0.038 , 0.288) | 10 ( 3 , 30 ) | 17 ( 15 , 19 ) | 9 ( 3 , 28 ) | 0.16 ( 0.05 , 0.31 ) | 0.26 ( 0.12 , 0.52 ) |
| **Haiti** | Gressier, Jacmel, Chabin | 0.209 ( 0.100 , 0.351 ) | 4 ( 3 , 8 ) | 10 ( 5 , 28 ) | 5 ( 3 , 9 ) | 0.11 ( 0.04 , 0.23 ) | 0.22 ( 0.11 , 0.42 ) |
| **India** | Chennai | 0.072 ( 0.024 , 0.172 ) | 21 (7 , 34 ) | NA | 14 ( 7 , 41 ) | 0.19 ( 0.06 , 0.35 ) | 0.21 ( 0.11 , 0.42 ) |
| **Singapore** | national | 0.034 ( 0.007 , 0.143 ) | NA | NA | 27 ( 9 , 177) | 0.18 ( 0.04 , 0.35 ) | 0.34 ( 0.15 , 0.66 ) |
| **Tanzania** | Zanzibar | 0.114 ( 0.034 , 0.254 ) | 7 ( 4 , 28 ) | 12 ( 8 , 18 ) | 7 (4 , 24 ) | 0.13 ( 0.03 , 0.29 ) | 0.32 ( 0.15 , 0.65 ) |
| **Venezuela** | Cana de Azucar | 0.059 ( 0.015 , 0.181 ) | 29 ( 6 , 40 ) | 99 ( 82 , 116 ) | 15 ( 6 , 64 ) | 0.17 ( 0.03 , 0.34 ) | 0.29 ( 0.13 , 0.59 ) |

**Table S9. Region-specific FOI estimates obtained under model A1.** Specifically, the table reports the FOI estimated under model C (normal likelihood) together with -the age at which we expect 50% seroprevalence and 70% seroprevalence and the average age at first infections.

| Country | Location | FOI | age_50%serop | age_70%serop | age_first_infection |
|---|---|---|---|---|---|
| **Brazil** | Fortaleza | 0.096 ( 0.08 , 0.114 ) | 7 ( 6 , 9 ) | 13 ( 11 , 15 ) | 11 ( 9 , 13 ) |
| **Mexico** | Pacific localities (Baja California, Baja California Sur, Nayarit, Sinloa, Sonora) | 0.053 ( 0.013 , 0.213 ) | 14 ( 4 , 45 ) | 25 ( 6 , 78 ) | 20 ( 5 , 65 ) |
| **Mexico** | South-Central localities (Guerrero, Morelos, Oaxaca, Puebla, Veracuz) | 0.061 ( 0.026 , 0.19 ) | 11 ( 3 , 21 ) | 19 ( 5 , 37 ) | 16 ( 4 , 31 ) |
| **Mexico** | South-East localities ( Campeche, Chiapas, Quintana Roo, Tabasco, Yucatan) | 0.118 ( 0.083 , 0.181 ) | 6 ( 4 , 9 ) | 10 ( 7 , 15 ) | 9 ( 6 , 13 ) |

**Table S9. Summary of FOI estimates identified in this review and in Cattarino et al [15].**

| Country | Location | Study date | FOI | Source |
|---|---|---|---|---|
| **Bangladesh** | blood service | 2022 | 0.133 | Review |
| **Australia** | Charters Towers | 1993 | 0.003 | Cattarino et al 2020 |
| **Bangladesh** | Dhaka city | 2012 | 0.045 | Review |
| **Bangladesh** | Dhaka City Corporation | 2012 | 0.014 | Cattarino et al 2020 |
| **Bangladesh** | NA | 2014-2016 | 0.002 | Cattarino et al 2020 |
| **Bangladesh** | NA | 2014-2016 | 0.002 | Cattarino et al 2020 |
| **Bangladesh** | NA | 2014-2016 | 0.001 | Cattarino et al 2020 |
| **Bangladesh** | NA | 2014-2016 | 0.003 | Cattarino et al 2020 |
| **Bangladesh** | NA | 2014-2016 | 0.004 | Cattarino et al 2020 |
| **Bangladesh** | NA | 2014-2016 | 0.042 | Cattarino et al 2020 |
| **Bangladesh** | NA | 2014-2016 | 0.001 | Cattarino et al 2020 |
| **Bangladesh** | NA | 2014-2016 | 0.031 | Cattarino et al 2020 |
| **Bangladesh** | NA | 2014-2016 | 0.002 | Cattarino et al 2020 |
| **Bangladesh** | NA | 2014-2016 | 0.003 | Cattarino et al 2020 |
| **Bangladesh** | NA | 2014-2016 | 0.002 | Cattarino et al 2020 |
| **Bangladesh** | NA | 2014-2016 | 0.001 | Cattarino et al 2020 |
| **Bangladesh** | NA | 2014-2016 | 0.003 | Cattarino et al 2020 |
| **Bangladesh** | NA | 2014-2016 | 0.001 | Cattarino et al 2020 |
| **Bangladesh** | NA | 2014-2016 | 0.001 | Cattarino et al 2020 |
| **Bangladesh** | NA | 2014-2016 | 0.001 | Cattarino et al 2020 |
| **Bangladesh** | NA | 2014-2016 | 0.001 | Cattarino et al 2020 |
| **Bangladesh** | NA | 2014-2016 | 0.002 | Cattarino et al 2020 |
| **Bangladesh** | NA | 2014-2016 | 0.002 | Cattarino et al 2020 |
| **Bangladesh** | NA | 2014-2016 | 0.002 | Cattarino et al 2020 |
| **Bangladesh** | NA | 2014-2016 | 0.002 | Cattarino et al 2020 |
| **Bangladesh** | NA | 2014-2016 | 0.003 | Cattarino et al 2020 |
| **Bangladesh** | NA | 2014-2016 | 0.033 | Cattarino et al 2020 |
| **Bangladesh** | NA | 2014-2016 | 0.005 | Cattarino et al 2020 |
| **Bangladesh** | NA | 2014-2016 | 0.006 | Cattarino et al 2020 |
| **Bangladesh** | NA | 2014-2016 | 0.004 | Cattarino et al 2020 |
| **Bangladesh** | NA | 2014-2016 | 0.009 | Cattarino et al 2020 |
| **Bangladesh** | NA | 2014-2016 | 0.001 | Cattarino et al 2020 |
| **Bangladesh** | NA | 2014-2016 | 0.000 | Cattarino et al 2020 |
| **Bangladesh** | NA | 2014-2016 | 0.001 | Cattarino et al 2020 |
| **Bangladesh** | NA | 2014-2016 | 0.001 | Cattarino et al 2020 |
| **Bangladesh** | NA | 2014-2016 | 0.001 | Cattarino et al 2020 |

| | | | | |
|---|---|---|---|---|
| **Bangladesh** | NA | 2014-2016 | 0.001 | Cattarino et al 2020 |
| **Bangladesh** | NA | 2014-2016 | 0.004 | Cattarino et al 2020 |
| **Bangladesh** | NA | 2014-2016 | 0.002 | Cattarino et al 2020 |
| **Bangladesh** | NA | 2014-2016 | 0.001 | Cattarino et al 2020 |
| **Bangladesh** | NA | 2014-2016 | 0.005 | Cattarino et al 2020 |
| **Bangladesh** | NA | 2014-2016 | 0.003 | Cattarino et al 2020 |
| **Bangladesh** | NA | 2014-2016 | 0.002 | Cattarino et al 2020 |
| **Bangladesh** | NA | 2014-2016 | 0.004 | Cattarino et al 2020 |
| **Bangladesh** | NA | 2014-2016 | 0.009 | Cattarino et al 2020 |
| **Bangladesh** | NA | 2014-2016 | 0.010 | Cattarino et al 2020 |
| **Bangladesh** | NA | 2014-2016 | 0.004 | Cattarino et al 2020 |
| **Bangladesh** | NA | 2014-2016 | 0.001 | Cattarino et al 2020 |
| **Bangladesh** | NA | 2014-2016 | 0.005 | Cattarino et al 2020 |
| **Bangladesh** | NA | 2014-2016 | 0.018 | Cattarino et al 2020 |
| **Bangladesh** | NA | 2014-2016 | 0.019 | Cattarino et al 2020 |
| **Bangladesh** | NA | 2014-2016 | 0.001 | Cattarino et al 2020 |
| **Bangladesh** | NA | 2014-2016 | 0.007 | Cattarino et al 2020 |
| **Bangladesh** | NA | 2014-2016 | 0.001 | Cattarino et al 2020 |
| **Bangladesh** | NA | 2014-2016 | 0.002 | Cattarino et al 2020 |
| **Bangladesh** | NA | 2014-2016 | 0.003 | Cattarino et al 2020 |
| **Bangladesh** | NA | 2014-2016 | 0.002 | Cattarino et al 2020 |
| **Bangladesh** | NA | 2014-2016 | 0.005 | Cattarino et al 2020 |
| **Bangladesh** | NA | 2014-2016 | 0.001 | Cattarino et al 2020 |
| **Bangladesh** | NA | 2014-2016 | 0.002 | Cattarino et al 2020 |
| **Bangladesh** | NA | 2014-2016 | 0.001 | Cattarino et al 2020 |
| **Bangladesh** | NA | 2014-2016 | 0.001 | Cattarino et al 2020 |
| **Bangladesh** | NA | 2014-2016 | 0.001 | Cattarino et al 2020 |
| **Bangladesh** | NA | 2014-2016 | 0.001 | Cattarino et al 2020 |
| **Bangladesh** | NA | 2014-2016 | 0.001 | Cattarino et al 2020 |
| **Bangladesh** | NA | 2014-2016 | 0.001 | Cattarino et al 2020 |
| **Bangladesh** | NA | 2014-2016 | 0.001 | Cattarino et al 2020 |
| **Bangladesh** | NA | 2014-2016 | 0.001 | Cattarino et al 2020 |
| **Bangladesh** | NA | 2014-2016 | 0.001 | Cattarino et al 2020 |
| **Bangladesh** | NA | 2014-2016 | 0.002 | Cattarino et al 2020 |
| **Bangladesh** | NA | 2014-2016 | 0.001 | Cattarino et al 2020 |
| **Bangladesh** | NA | 2014-2016 | 0.001 | Cattarino et al 2020 |
| **Bangladesh** | NA | 2014-2016 | 0.000 | Cattarino et al 2020 |
| **Bangladesh** | NA | 2014-2016 | 0.000 | Cattarino et al 2020 |
| **Brazil** | Sau Paulo | 2015-2016 | 0.030 | Review |

| | | | | |
|---|---|---|---|---|
| **Brazil** | Fortaleza | 2011-2015 | 0.096 | Review |
| **Brazil** | Recife, State of Pernambuco | 2005_2006 | 0.014 | Cattarino et al 2020 |
| **Brazil** | Niterói, State of Rio de Janeiro | 1991 | 0.026 | Cattarino et al 2020 |
| **Brazil** | Paracambi, State of Rio de Janeiro | 1994 | 0.013 | Cattarino et al 2020 |
| **Brazil** | Acrelândia, State of Acre | 2004_2005 | 0.002 | Cattarino et al 2020 |
| **Brazil** | São Luis Island, State of Maranhão | 1996 | 0.003 | Cattarino et al 2020 |
| **Brazil** | Campinas, State of São Paulo | 1998 | 0.002 | Cattarino et al 2020 |
| **Burkina Faso** | Ougadougou | 2015-2017 | 0.063 | Review |
| **Cameroon** | Garoua | 2006 | 0.002 | Cattarino et al 2020 |
| **Cameroon** | Douala | 2006 | 0.009 | Cattarino et al 2020 |
| **Cameroon** | Yaounde | 2006 | 0.001 | Cattarino et al 2020 |
| **Colombia** | Anapoima, Apulo, Buenaventura, Quibdo, Tumaco,Tierralta | 2013-2015 | 0.047 | Review |
| **Colombia** | Quobdo | 2015* | 0.041 | Review |
| **Colombia** | Santander | 2015* | 0.029 | Cattarino et al 2020 |
| **Costa Rica** | San José | 2002_2003 | 0.028 | Cattarino et al 2020 |
| **Cuba** | Santiago de Cuba | 1997_1998 | 0.004 | Cattarino et al 2020 |
| **Cuba** | El Cerro | 1983 | 0.010 | Cattarino et al 2020 |
| **Dominican Republic** | Santo Domingo district, Santo Domingo | 2002 | 0.029 | Cattarino et al 2020 |
| **Ecuador** | Town of Quininde | 2005-2009 | 0.094 | Review |
| **El Salvador** | Aguilares | 2000_2001 | 0.031 | Cattarino et al 2020 |
| **France (Carribean)** | Guadeloupe and Martinique | 2011 | 0.078 | Review |
| **Haiti** | Gressier, Jacmel, Chabin | 2013 | 0.209 | Review |
| **India** | New Delhi, Hyderabad, Kalyani, Wardha, Mumbai, Bangalore | 2012 | 0.127 | Review |
| **India** | Pradesh, Tripura, Meghalaya, Assam, Bihar, West Bengal, Odisha, Rajasthan, Madhya Pradesh, Maharashtra, Andhra Pradesh, Karnataka, Tamil Nadu | 2017 | 0.046 | Review |
| **India** | Chennai | 2011 | 0.072 | Review |
| **India** | Delhi | 2012 | 0.043 | Review |
| **India** | Denpasar Bali | 2020-2021 | 0.036 | Review |
| **India** | Pune | 2017 | 0.078 | Review |
| **India** | Vadu area | 2011 | 0.021 | Review |
| **India** | Andaman Islands | 1988_1989 | 0.001 | Cattarino et al 2020 |
| **India** | Chennai | 2011 | 0.040 | Cattarino et al 2020 |
| **India** | New Delhi | 2011-2012 | 0.030 | Cattarino et al 2020 |
| **India** | New Delhi | 2011-2012 | 0.035 | Cattarino et al 2020 |
| **India** | Kalyani | 2011-2012 | 0.010 | Cattarino et al 2020 |
| **India** | Wardha | 2011-2012 | 0.038 | Cattarino et al 2020 |
| **India** | Mumbai | 2011-2012 | 0.051 | Cattarino et al 2020 |

| | | | | |
|---|---|---|---|---|
| **India** | Medchal | 2011-2012 | 0.029 | Cattarino et al 2020 |
| **India** | Medchal | 2011-2012 | 0.029 | Cattarino et al 2020 |
| **India** | Bangalore | 2011-2012 | 0.032 | Cattarino et al 2020 |
| **India** | Pune, Koregaon Bhima | 2011 | 0.011 | Cattarino et al 2020 |
| **India** | Pune, Pimpale Jagatap | 2012 | 0.006 | Cattarino et al 2020 |
| **Indonesia** | national | 2014 | 0.144 | Review |
| **Indonesia** | national | 2014 | 0.149 | Review |
| **Indonesia** | Gondokusuman subdistrict, Yogyakarta province | 1995 | 0.030 | Cattarino et al 2020 |
| **Indonesia** | Jakarta | 2014 | 0.037 | Cattarino et al 2020 |
| **Kenya** | national | 2007 | 0.004 | Review |
| **Kenya** | Kenya | 2007 | 0.002 | Cattarino et al 2020 |
| **Kenya** | Kwale County | 2009-2011 | 0.006 | Cattarino et al 2020 |
| **Kenya** | Busia | 2004 | 0.000 | Cattarino et al 2020 |
| **Kenya** | Malindi | 2004 | 0.003 | Cattarino et al 2020 |
| **Kenya** | Maralal | 2004 | 0.000 | Cattarino et al 2020 |
| **Laos** | Khammouane | 2007_2008 | 0.005 | Cattarino et al 2020 |
| **Laos** | Vientiane | 2006 | 0.010 | Cattarino et al 2020 |
| **Malaysia** | Federal territory of Kuala Lumpur, Perak, Kedah, Penang, Johor, Pahang, Kelantan and Sabah | 2008-2009 | 0.010 | Review |
| **Malaysia** | Damansara Damai | 2018-2019 | 0.033 | Review |
| **Malaysia** | Peninsular Malaysia | 2006-2012 | 0.038 | Review |
| **Malaysia** | Petaling district | 2018 | 0.059 | Review |
| **Malaysia** | Sungai Segamat | 2015 | 0.029 | Review |
| **Malaysia** | Segamat district, Johor state | 2015 | 0.008 | Cattarino et al 2020 |
| **Mayotte** | Mayotte | 2006 | 0.003 | Cattarino et al 2020 |
| **Mexico** | Pacific localities (Baja California, Nayarit, Sinloa, Sonora) | 2016 | 0.053 | Review |
| **Mexico** | South-Central localities (Guerrero,Morelos, Oaxaca, Puebla, Veracuz) | 2016 | 0.061 | Review |
| **Mexico** | South-East localities (Campeche, Chiapas, Quintana Roo, Tabasco, Yucatan) | 2016 | 0.118 | Review |
| **Mexico** | Yucatan | 2014 | 0.045 | Review |
| **Mexico** | State of Morelos | 2011 | 0.044 | Review |
| **Mexico** | Matamoros, Tamaulipas | 2004 | 0.009 | Cattarino et al 2020 |
| **Mexico** | Matamoros, Tamaulipas | 2005 | 0.009 | Cattarino et al 2020 |
| **Mexico** | Morelos | 2015* | 0.014 | Cattarino et al 2020 |
| **Namibia** | Windhoek | 2011-2012 | 0.001 | Cattarino et al 2020 |
| **Nigeria** | Kainji Lake | 1980 | 0.006 | Cattarino et al 2020 |
| **Pakistan** | Lahore | 2010-2015 | 0.060 | Review |
| **Pakistan** | Khyber Pakhtunkhawa | pre_2013 | 0.002 | Cattarino et al 2020 |

| | | | | |
|---|---|---|---|---|
| **Pakistan** | Lahore | 2012 | 0.008 | Cattarino et al 2020 |
| **Pakistan** | Lahore | 2011-2016 | 0.020 | Cattarino et al 2020 |
| **Papua New Guinea** | Madang province | 2007_2008 | 0.056 | Cattarino et al 2020 |
| **Peru** | Iquitos | 1992 | 0.009 | Cattarino et al 2020 |
| **Peru** | Santa Clara, Iquitos | 1996 | 0.003 | Cattarino et al 2020 |
| **Peru** | Iquitos | 1999 | 0.032 | Cattarino et al 2020 |
| **Philippines** | Cebu | 2015* | 0.061 | Cattarino et al 2020 |
| **Reunion** | Reunion Island | 2008 | 0.001 | Cattarino et al 2020 |
| **Saudi Arabia** | Makkah, Madinah,Jeddah, Jizan | 2016-2017 | 0.008 | Review |
| **Saudi Arabia** | Jeddah | 2015* | 0.017 | Review |
| **Saudi Arabia** | Jeddah | pre_2016 | 0.005 | Cattarino et al 2020 |
| **Singapore** | national | 2008-2010 | 0.020 | Review |
| **Singapore** | blood service | 2009-2010 | 0.049 | Review |
| **Singapore** | national | 2010 | 0.034 | Review |
| **Singapore** | Singapore | 2010 | 0.006 | Cattarino et al 2020 |
| **Singapore** | Singapore | 1982_1984 | 0.014 | Cattarino et al 2020 |
| **Singapore** | Singapore | 2007 | 0.007 | Cattarino et al 2020 |
| **Singapore** | Singapore | 2004_2007 | 0.006 | Cattarino et al 2020 |
| **Singapore** | Singapore | 2009-2010 | 0.005 | Cattarino et al 2020 |
| **Sri Lanka** | City of Colombo | 2008-2010 | 0.149 | Review |
| **Sri Lanka** | Maharagama Medical Officer Health, Colombo | pre_2006 | 0.010 | Cattarino et al 2020 |
| **Sri Lanka** | Colombo | 2008_2009 | 0.032 | Cattarino et al 2020 |
| **Sri Lanka** | Colombo | 2008 | 0.034 | Cattarino et al 2020 |
| **Sudan** | Karima | 1989 | 0.002 | Cattarino et al 2020 |
| **Sudan** | Kassala state | 2011 | 0.001 | Cattarino et al 2020 |
| **Taiwan** | Taipei, Taoyuan, Tainan | 2010 | 0.005 | Review |
| **Taiwan** | Nanzih district, Kaohsiung City | 2015-2016 | 0.001 | Review |
| **Taiwan** | Sanmin district, Kaohsiung City | 2015-2016 | 0.001 | Review |
| **Taiwan** | Pingtung County | 1997 | 0.005 | Cattarino et al 2020 |
| **Tanzania** | Buhigwe, Kalambo, Kilindi, Kinondoni, Kondoa, Kyela, Mvomero and Ukerewe | 2018 | 0.011 | Review |
| **Tanzania** | Zanzibar | 2010 | 0.114 | Review |
| **Tanzania** | Zanzibar | 2011 | 0.007 | Cattarino et al 2020 |
| **Thailand** | Mukdahn, Ubon, Ratchathani, Savannakhet and Champasak | 2019 | 0.083 | Review |
| **Thailand** | Ayutthaya, Lop Buri, Narathiwat and Trag | 2014 | 0.070 | Review |
| **Thailand** | Ratchaburi province | 2012-2015 | 0.136 | Review |
| **Thailand** | Ratchaburi province | 2019-2020 | 0.094 | Review |
| **Thailand** | Bangkok | 2000 | 0.028 | Cattarino et al 2020 |
| **Thailand** | Mueang district, Ratchaburi Province | 2000 | 0.035 | Cattarino et al 2020 |
| **Thailand** | Mueang Rayong District | 2010 | 0.019 | Cattarino et al 2020 |

| | | | | |
|---|---|---|---|---|
| **Thailand** | Rayong, Rayong Province | 1980 | 0.044 | Cattarino et al 2020 |
| **Thailand** | Ayutthaya province | 2014 | 0.021 | Cattarino et al 2020 |
| **Thailand** | Narathiwat province | 2014 | 0.071 | Cattarino et al 2020 |
| **United States** | Brownsville, Texas | 2004 | 0.003 | Cattarino et al 2020 |
| **United States** | Brownsville, Texas | 2005 | 0.003 | Cattarino et al 2020 |
| **Venezuela** | Cana de Azucar | 2010-2011 | 0.059 | Review |
| **Venezuela** | Isla de San Carlos | 1995 | 0.005 | Cattarino et al 2020 |
| **Venezuela** | Maracay | 2010 | 0.025 | Cattarino et al 2020 |
| **Vietnam** | Cao Lanh City, Dong Thap Province | 1996_1997 | 0.036 | Cattarino et al 2020 |
| **Vietnam** | Binh Thuan Province | pre_2005 | 0.028 | Cattarino et al 2020 |

*The date of the survey was not specified, instead we report the year of publication of the article.

### Figures

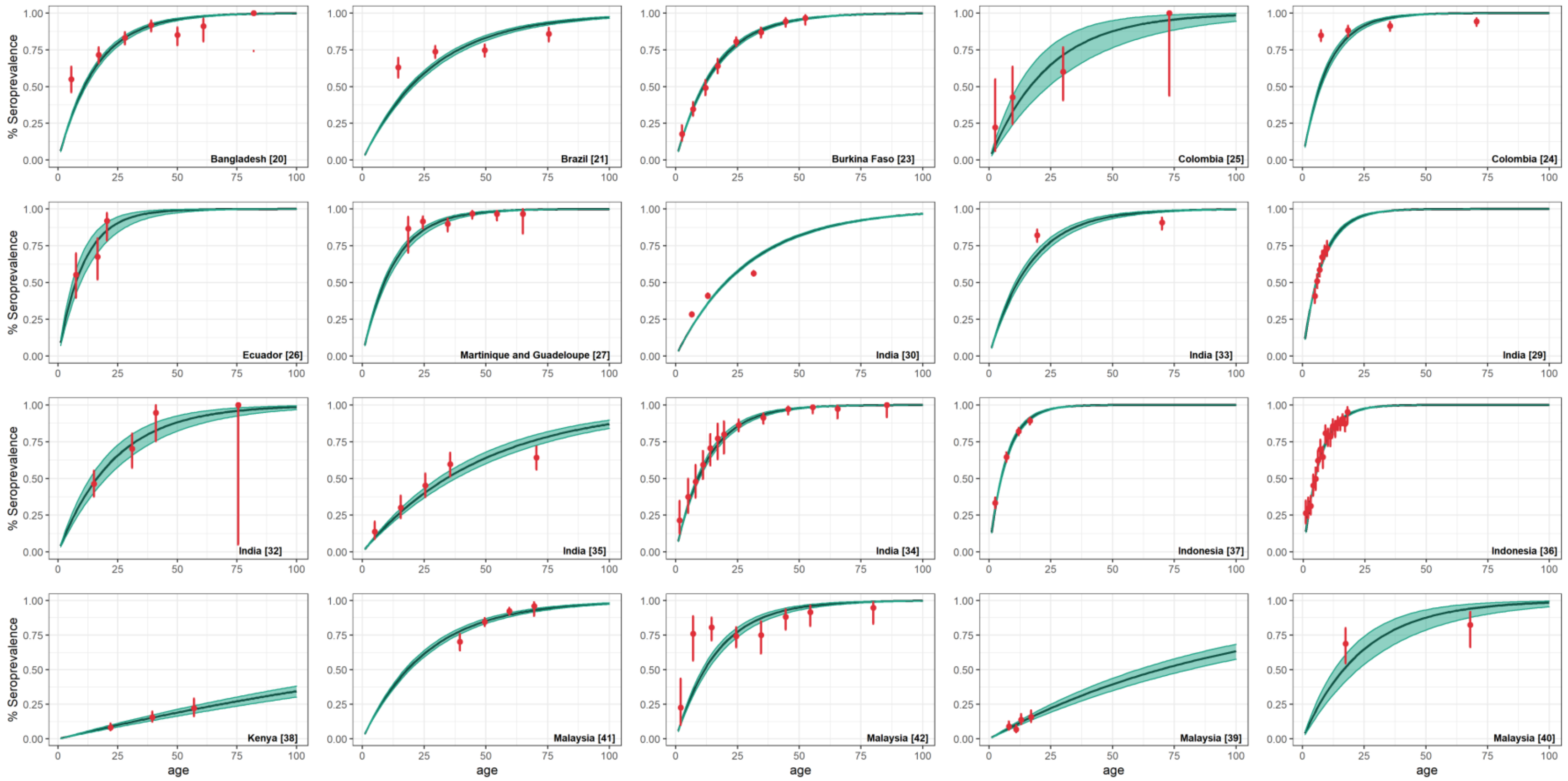


**Figure S1 Model fit obtained under model A1 (binomial) part 1.** The points (in red) represent all available samples with their exact binomial 95% confidence interval (CI) and the continuous black line and green shading represent the median and 95% credible interval (CrI) obtained from 1000 random samples of the estimated FOI from the posterior distribution.

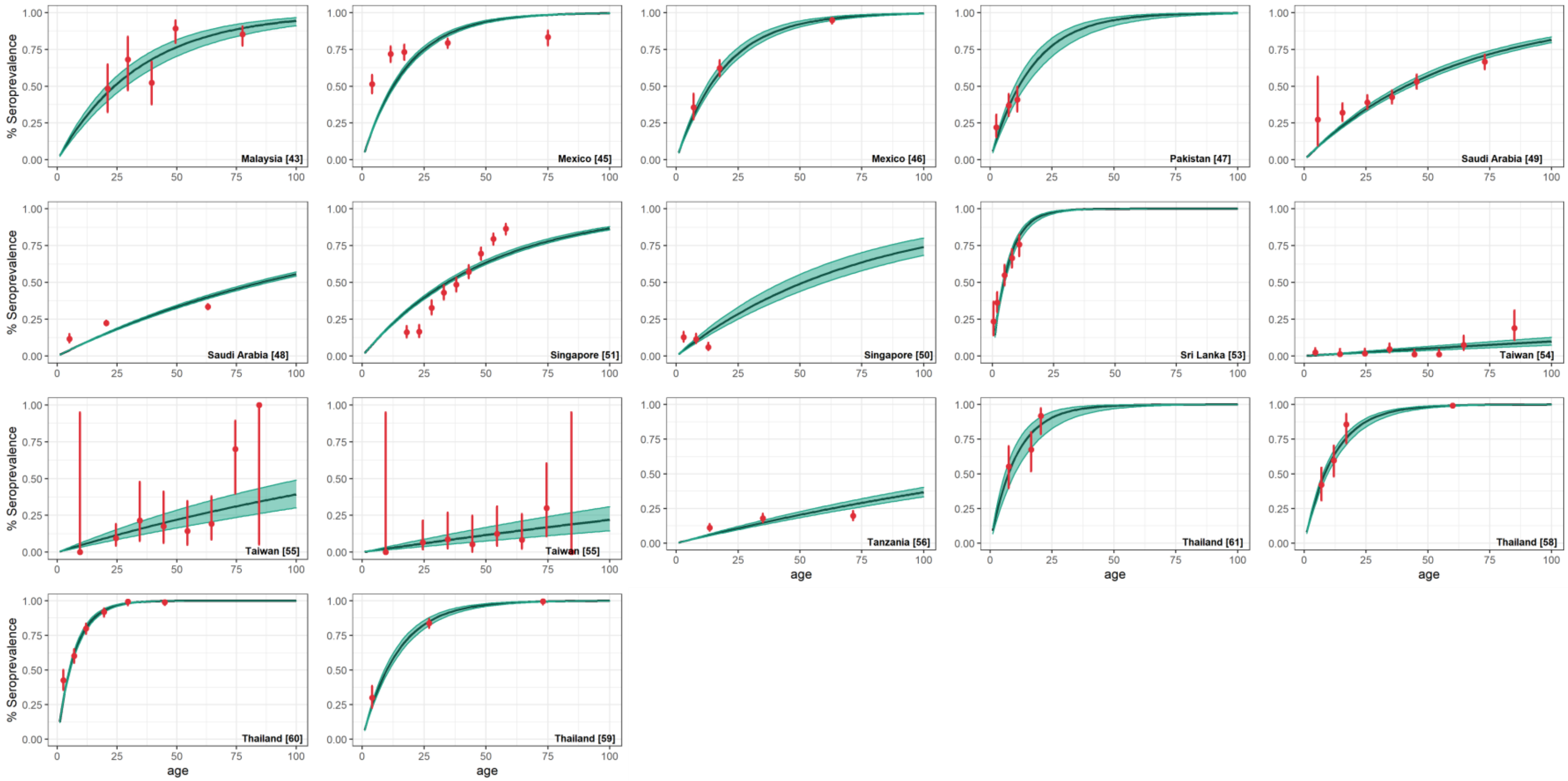


**Figure S2 Model fit obtained under model A1 (binomial) part 2.** The points (in red) represent all available samples with their exact binomial 95% confidence interval (CI) and the continuous black line and green shading represent the median and 95% credible interval (CrI) obtained from 1000 random samples of the estimated FOI from the posterior distribution.

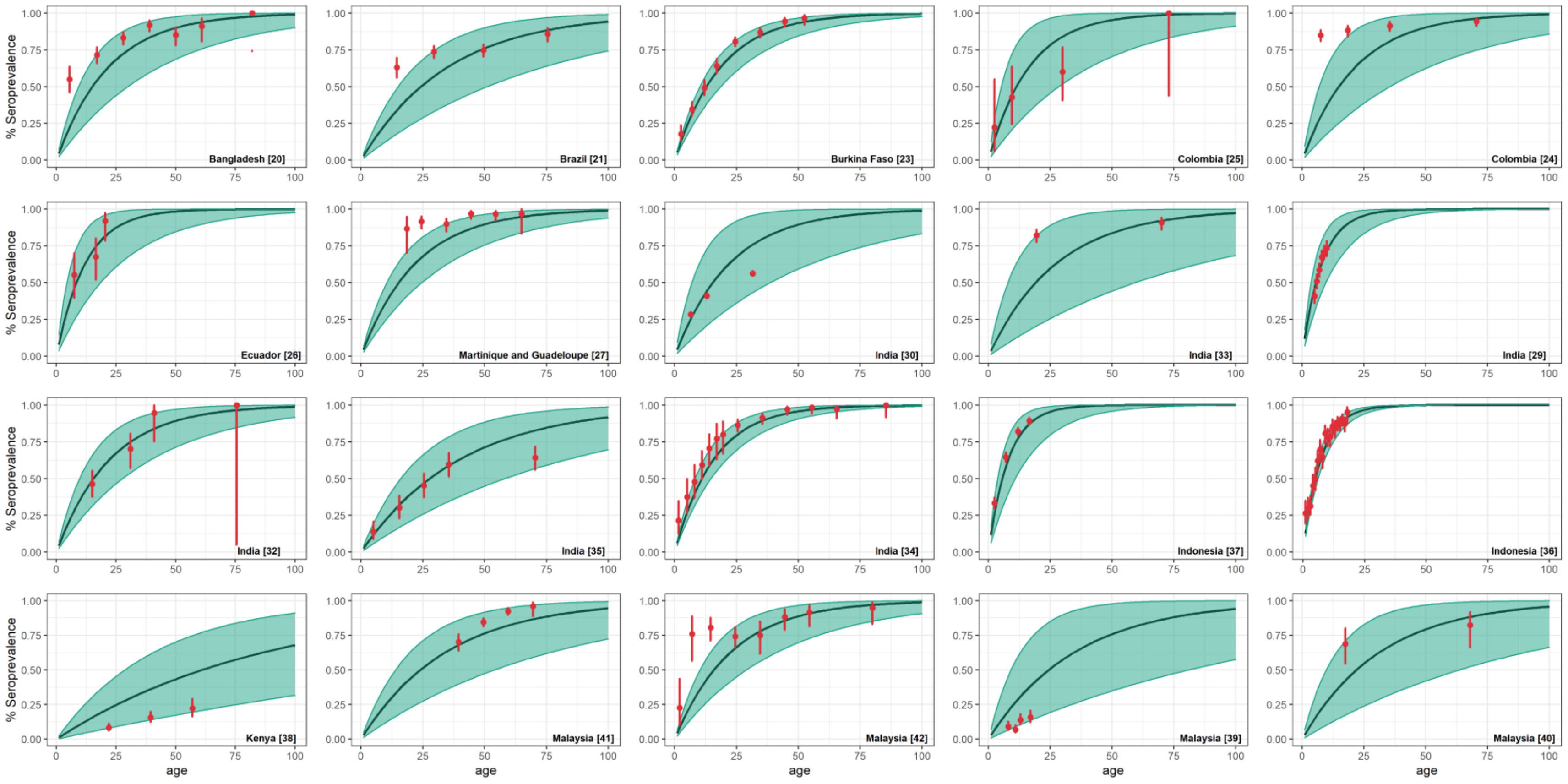


**Figure S3 Model fit obtained under model A2 (betabinomial) part 1.** The points (in red) represent all available samples with their exact binomial 95% confidence interval (CI) and the continuous black line and green shading represent the median and 95% credible interval (CrI) obtained from 1000 random samples of the estimated FOI from the posterior distribution.

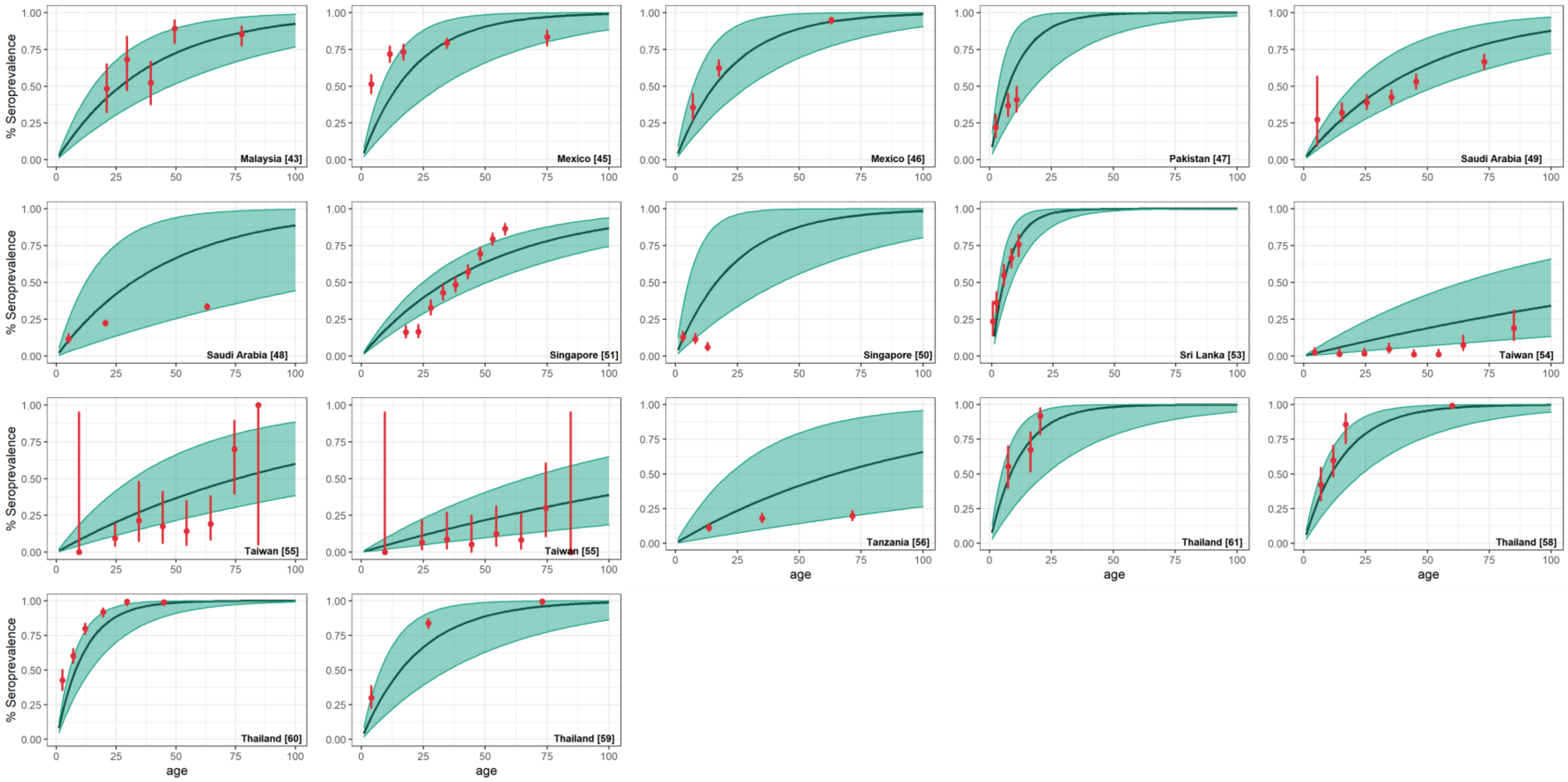


**Figure S4 Model fit obtained under model A2 (betabinomial) part 2.** The points (in red) represent all available samples with their exact binomial 95% confidence interval (CI) and the continuous black line and green shading represent the median and 95% credible interval (CrI) obtained from 1000 random samples of the estimated FOI from the posterior distribution.

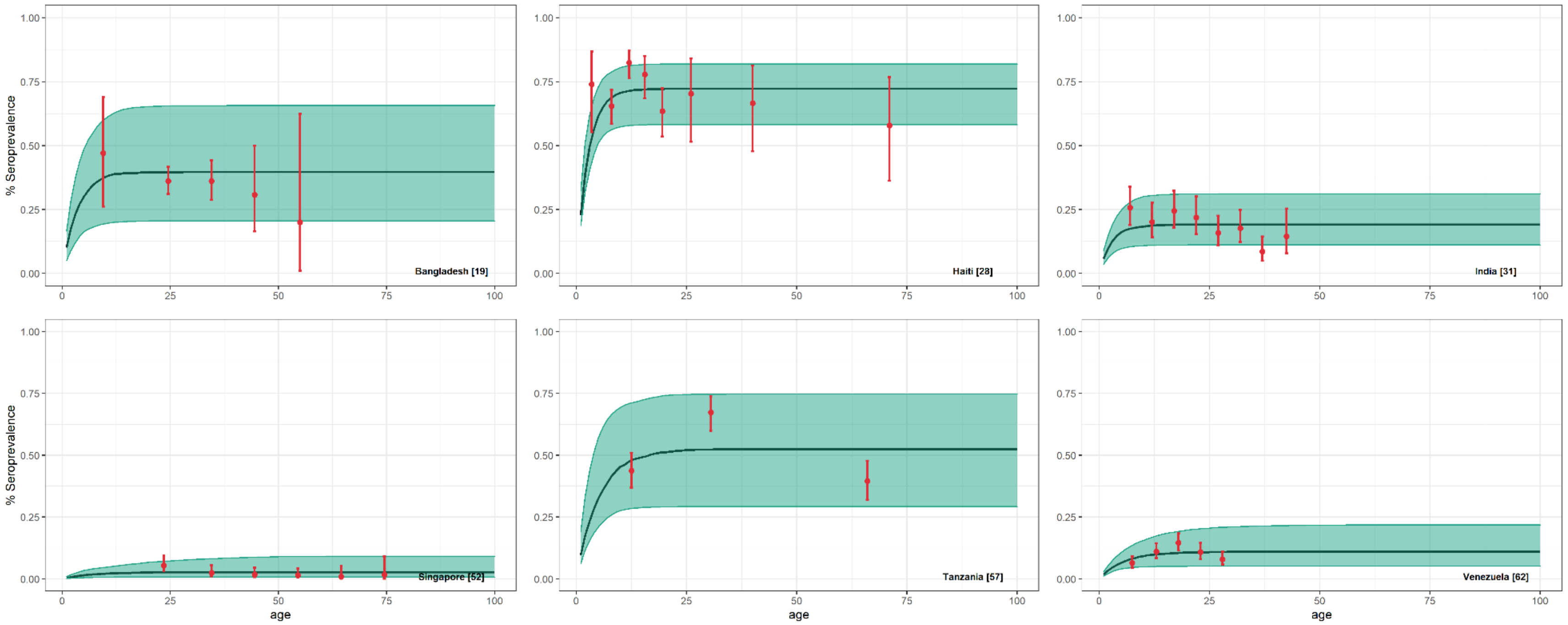


**Figure S5 Model fit obtained under model B1 (binomial).** The points (in red) represent all available samples with their exact binomial 95% confidence interval (CI) and the continuous black line and green shading represent the median and 95% credible interval (CrI) obtained from 1000 random samples of the estimated FOI from the posterior distribution.

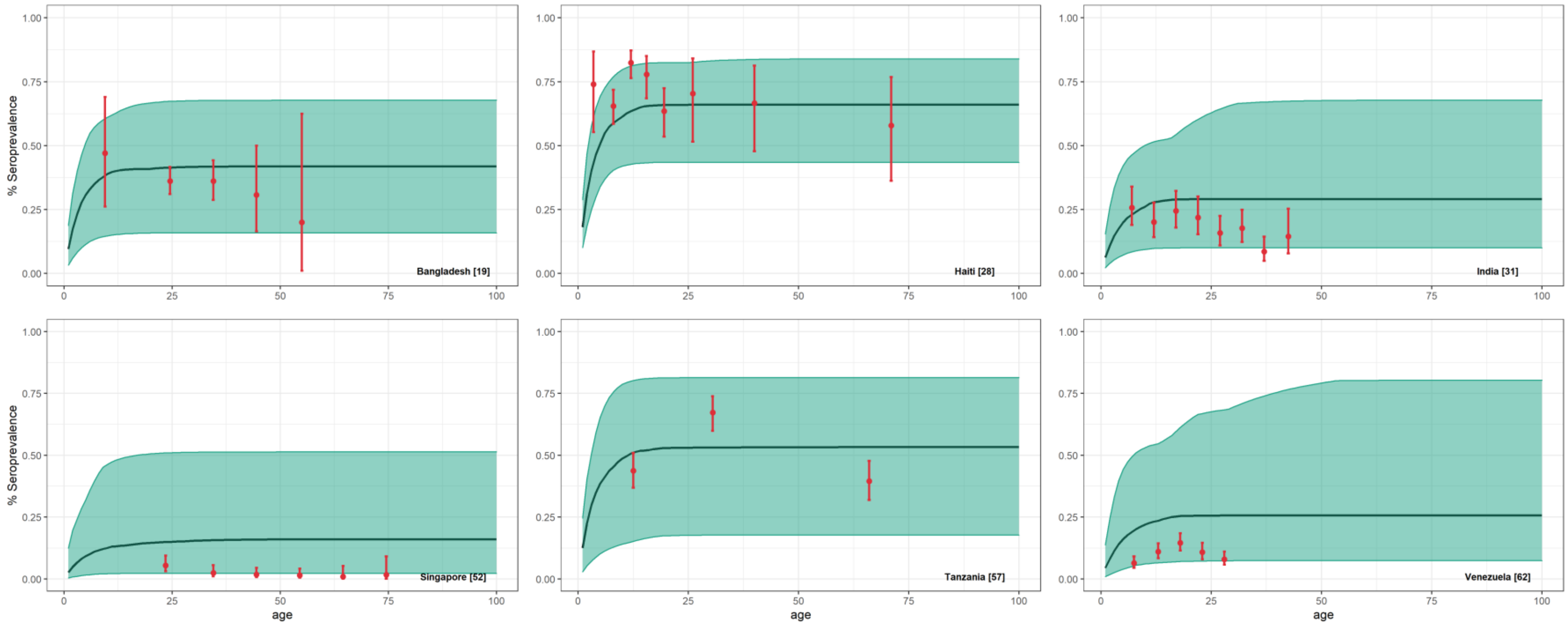


**Figure S6 Model fit obtained under model B2 (betabinomial).** The points (in red) represent all available samples with their exact binomial 95% confidence interval (CI) and the continuous black line and green shading represent the median and 95% credible interval (CrI) obtained from 1000 random samples of the estimated FOI from the posterior distribution.

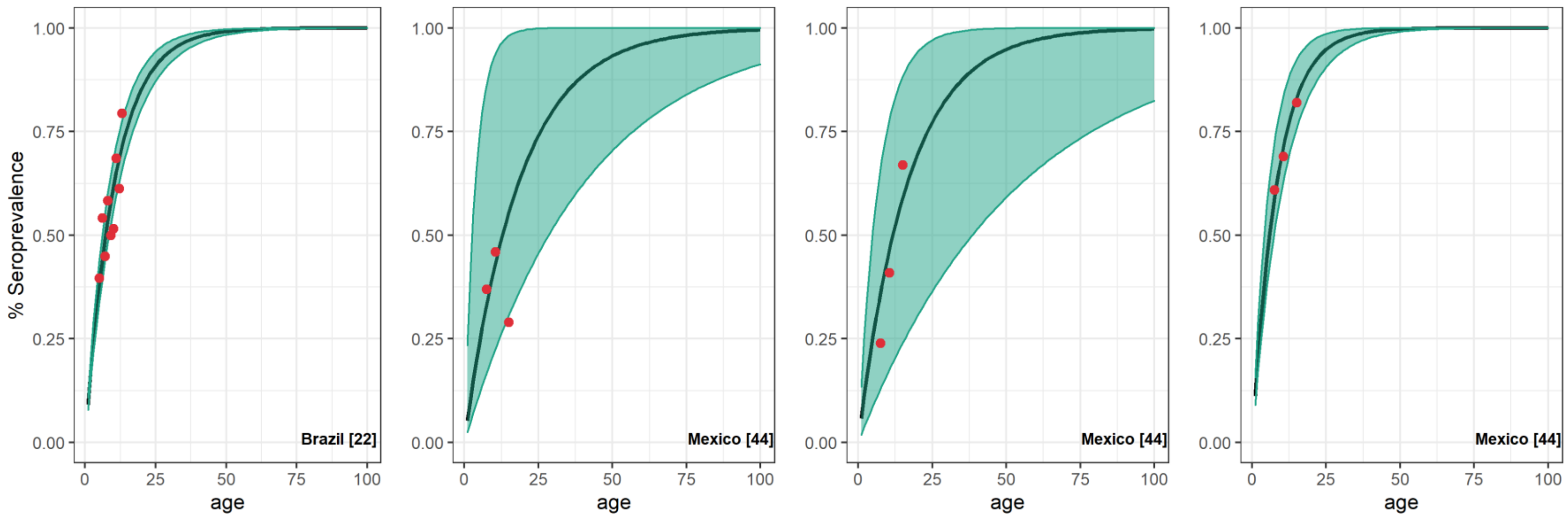


**Figure S7 Model fit obtained under model C (normal).** The points (in red) represent all available samples with their exact binomial 95% confidence interval (CI) and the continuous black line and green shading represent the median and 95% credible interval (CrI) obtained from 1000 random samples of the estimated FOI from the posterior distribution.

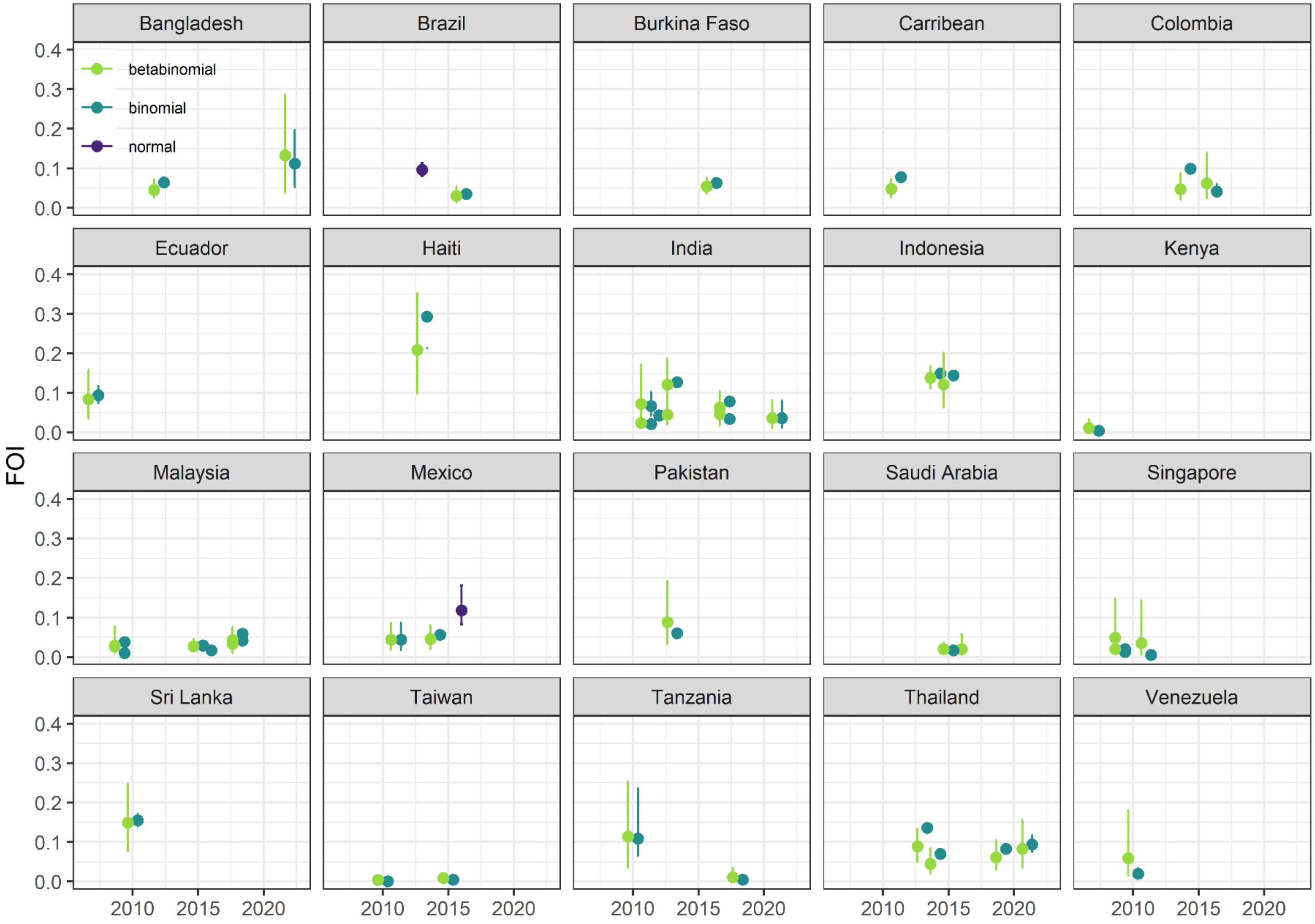


**Figure S8 FOI estimates obtained under models A1, A2, B1, B2 and C for each dataset across several countries.** The x axis reports the date of the serological survey. Each FOI estimate is reported as median and 95% credible interval (CrI). The results from the model A1 and B1 (binomial likelihood) are reported in light blue and compared with those from model A2 and B2 (betabinomial likelihood) highlighted in green. FOI estimates from model C (normal likelihood) are reported in purple.
